# Brief psychological interventions to improve mental health outcomes in refugee populations: A systematic review

**DOI:** 10.1101/2024.03.08.24303974

**Authors:** Nadia A. Daniel, Xin Liu, Elizabeth T. Thomas, Emily Eraneva-Dibb, Al-Maz Ahmad, Carl Heneghan

## Abstract

**Background:** Refugees, asylum seekers, and internally displaced people experience a high burden of mental health problems owing to their experiencing traumas and stressful events.

**Objective:** To summarise the available evidence and analyse the efficacy of brief psychological interventions (< 3 months) on improving mental health outcomes, including depression, anxiety, and post-traumatic stress disorder (PTSD)-related symptoms in refugees.

**Method:** We searched Medline, EMBASE, PsycINFO, CINAHL, and Global Index Medicus from inception to 19 December 2023. We included controlled studies using any cognitive behavioural therapy (CBT) or CBT-based therapies delivered over a short time (< 3 months), which reported mental health outcomes pre-and post-intervention. We conducted meta-analyses using random effects to derive pooled summary statistics.

**Results:** 34 eligible studies across 36 publications were retrieved for analysis, and 33 studies were included in the meta-analysis. There was an overall improvement in immediate mental health outcomes for all three domains, with analysis of 13 studies on anxiety outcomes (SMD −1.12, 95% CI −1.72 to −0.52), 20 studies on depression (SMD −1.04, 95% CI −1.97 to −0.11), and 24 studies on PTSD (SMD −0.82, 95% CI −1.20 to −0.45). At 3 to 6-month follow-up, however, analysis of mental health outcomes shows no significant change from baseline, with standard mean differences of 0.24 (95% CI −0.94 to 1.42) across 4 studies, −0.73 (95% CI −2.14 to 0.68) across 9 studies, and 0.29 (95% CI −0.94 to 1.53) across 12 studies for anxiety, depression, and PTSD respectively.

**Conclusion:** Low-level evidence shows brief psychological interventions positively affect refugees and internally displaced people’s mental well-being. Heterogeneity was high, even among subgroups, impacting our findings’ generalisability.

**HIGHLIGHTS:**

- We analysed the evidence on the use of brief CBT-based psychological interventions to improve mental health outcomes in forcibly displaced persons.
- These interventions had a positive effect on anxiety, depression, and PTSD, though there was high heterogeneity between studies.
- Positive effects on mental health disappeared at long-term follow-up.

## INTRODUCTION

The population of refugees, asylum seekers, and internally displaced people has grown to 110 million in 2023^1^. The traumatic issues that force people to leave their homes, including persecution, violence, and human rights violations, have profound impacts on mental health^2–4^. They are then likely to experience additional stressful events during migration and resettlement in a new country ^4–6^. As a result, forcibly displaced people have a markedly high prevalence of mental illness, notably posttraumatic stress disorder (PTSD), depression, and anxiety^7–10^.

Psychosocial interventions have proven to be effective in both preventing and treating mental health problems. For asylum seekers and refugees, cognitive behavioural therapy (CBT) and narrative exposure therapy (NET) have been shown to be effective in relieving symptoms of PTSD, depression, and anxiety^11,12^. CBT is a structured, goal-orientated form of therapy that aims to identify and modify harmful patterns of thinking and behaviour ^13^ NET is short-term, trauma-focused CBT whereby therapist and patient work to create a timeline of the patient’s life, rebuilding the autobiographical memory, allowing for the reduction of anxiety^14^.

These interventions can benefit forcibly displaced people; however, they are often challenging to implement due to the short-term nature of their living situation (such as refugee camps)^15,16^, or limited resources in host countries, with 75 percent of forcibly displaced people hosted by low- and middle-income countries^17^.

A possible solution to these challenges is brief psychological interventions where a restricted number of sessions are delivered over a short-term period. These are less resource intensive, particularly if delivered by trained lay workers rather than mental health specialists, and more appropriate for transient settings such as refugee camps. Examples include the low-intensity, transdiagnostic interventions developed by the World Health Organisation (WHO), including Problem Management Plus (PM+), which uses aspects of CBT to improve one’s management of practical problems and associated mental health issues^18^ and Self-Help Plus (SH+), a guided self-help program delivered through a pre-recorded audio course, based on acceptance and commitment therapy (a form of CBT)^19^.

In 2020, a Cochrane scoping review^20^ found a gap in the evidence on mental health interventions for refugees, asylum seekers, and internally displaced persons. There have been several recent systematic reviews looking at psychological interventions for refugees and asylum seekers. Two focus only on low-intensity, transdiagnostic interventions ^21,22^; one on psychological interventions in children ^23^; and one on psychosocial interventions for PTSD only in low- and middle-income countries ^24^. To date, no systematic review has examined the effect of brief CBT-based interventions on varied outcomes of mental health symptoms.

Therefore, to address this gap, we aimed to synthesise the body of evidence on short-term CBT-based interventions that were delivered in less than 3 months. The timing reflects the transient nature of housing situations for many forcibly displaced persons and the limited resources for their mental health provision.

## METHODS

This systematic review adheres to standards outlined by Cochrane Collaboration^25^ and is reported according to the Preferred Reporting Items for Systematic Reviews and Meta-Analyses (PRISMA^24^). Differences between the preregistration of this review (preregistration-ID: 10.17605/OSF.IO/9CXU4) and the final review are presented in the Supplementary Material (**SM1**).

### Search strategy

We developed the search strategy based on previous studies^12,20^ and consulted an information specialist to assess the search keywords and give recommendations for the final search strategy.

We searched databases from inception, with the search last updated on December 19, 2023. We searched five databases: Ovid Medline, Ovid EMBASE, Ovid PsycINFO, CINAHL, and Global Index Medicus. Further searches on Google Scholar and the ICTRP (International Clinical Trials Registry Platform) were conducted to ensure all relevant comprehensiveness of the search strategy.

Full search strategies are presented in **SM2**. We also screened the reference lists of included studies and related systematic reviews for inclusion.

### Selection criteria

We included controlled trials using any CBT (or CBT-based therapies) delivered over a short period of time, as defined by the study, or as interventions that took place in a time frame of up to 3 months, reporting mental health outcomes pre-and post-intervention. We excluded observational studies and feasibility trials without a control group, case reports, case series and reviews of internet-delivered CBT interventions.

We used 3 months as the specific time limit because studies which describe their interventions as ‘brief’ generally take place within this timeframe^26–28^. We focussed on CBT-based interventions, as these interventions are feasible and generalisable in forcibly displaced persons and have evidence to support their use^12,29,30^.

The review aimed to:

1. demonstrate if these interventions are efficacious in improving a range of mental health symptoms, particularly PTSD, anxiety, or depression,
2. assess if improvements can be sustained in the long term; and
3. identify factors that influence the effectiveness of the psychological interventions.

### Study selection

All abstracts and full texts retrieved from our search strategy were screened in duplicate by three authors (ND, LX, EED), and any discrepancies were resolved by discussion with a third author.

### Data extraction

Three investigators extracted data in duplicate (ND, LX, EED) using a standardised data collection template with predefined data fields, including study characteristics, participant demographics, primary and secondary outcome measures, and drop-out rate. A third investigator resolved any conflicts.

### Quality appraisal

Three authors (ND, LX, EED) independently evaluated the risk of bias using the Cochrane Risk of Bias (RoB 2) tool^25^ for randomised trials and the ROBINS-I tool for non-randomised studies^26^. Any conflicts in the duplicated data were resolved by discussion with a third reviewer. We examined publication bias using visual inspections of funnel plots (reference) and Egger’s test.

### Data synthesis

As our primary outcome measure is the change in mental health outcomes as measured within 3 months of the intervention, the means for pre- and post-intervention measures were pooled, along with their standard deviations (SDs) and number of participants per study. We estimated standardised mean differences of anxiety, depression, and PTSD measures using Cohen’s d and Hedge’s g. Summary statistics for all trials were computed and displayed as forest plots using R version 4.3.2, using the meta package. We performed a random-effects meta-analysis for brief CBT-based psychological interventions in treating anxiety and depression and for brief CBT-based psychological interventions in treating PTSD.

We investigated the long-term effect of brief CBT-based interventions by performing post-hoc random-effects meta-analysis on mental health outcomes assessed at 3-6 months and 7-12 months post-intervention.

We examined heterogeneity by performing post-hoc subgroup analyses, grouping studies by the type of mental health assessment tools used, intervention type, population type, personnel conducting the intervention, sample size (>100 or <100), and whether the setting was in a low or high-income country.

Finally, we performed sensitivity analysis by removing studies with a moderate and high risk of bias to assess the robustness of the outcome

## RESULTS

### Search outcomes

The search strategy yielded 4784 hits. After de-duplication and full-text screening, 33 studies (31 unique trials) were identified. Screening the reference lists of the identified studies yielded three further studies, which were included in the subsequent analysis (**Figure 1**).

**Figure 1.**
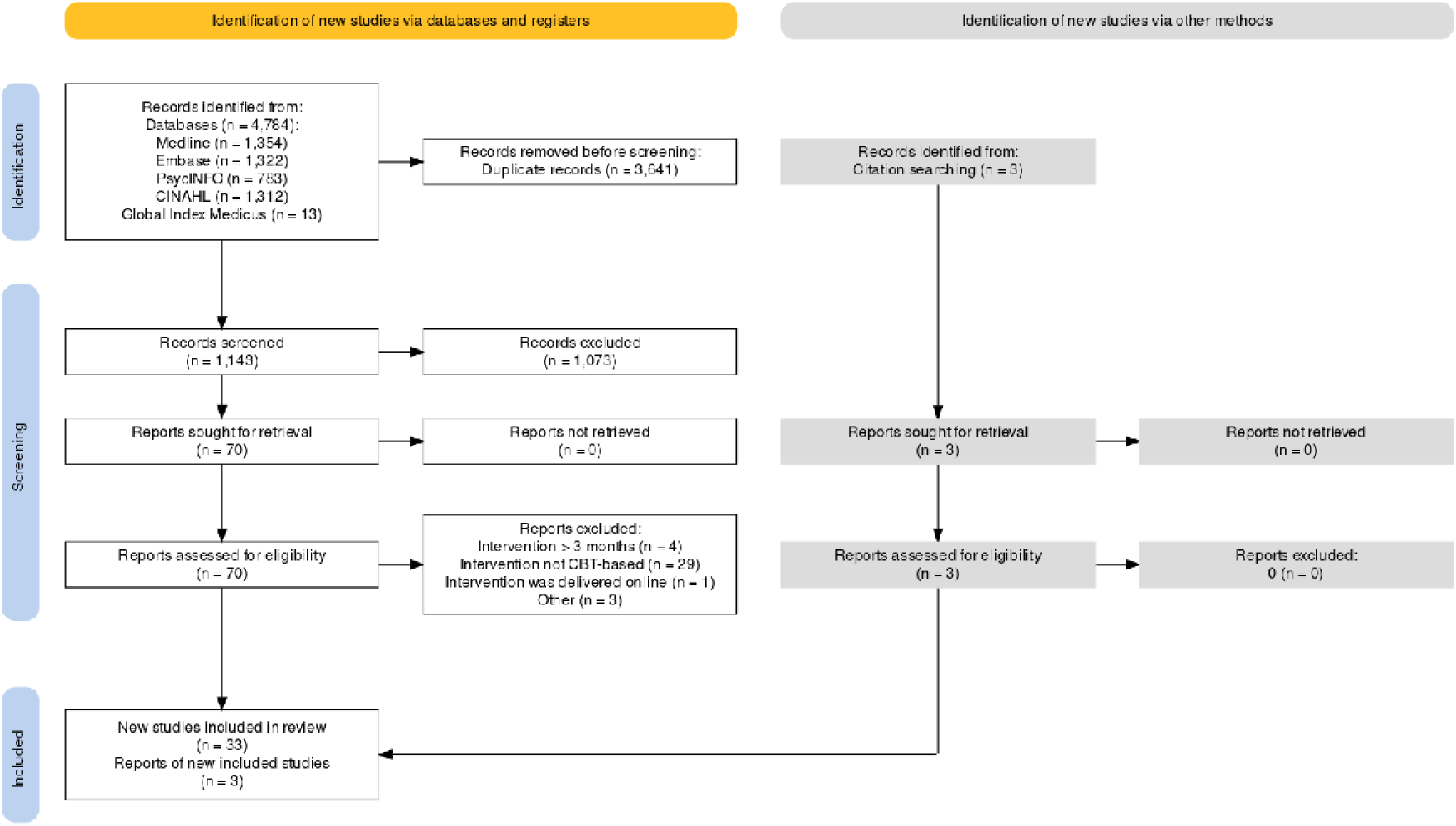
PRISMA flowchart of the study selection process.

### Study characteristics

**Tables 1 & 2** report the characteristics of the 36 included studies and their extracted outcomes. Of these studies, 27 were carried out in adults, and the remaining nine were conducted in child and adolescent populations. Most studies were performed in Germany and Uganda (5 studies each), Jordan, Turkey, and the USA (3 studies each). Only two trials involved internally displaced persons; the rest were conducted on refugees and/or asylum seekers. Most studies were RCTs (*n*=30, 83.3%).

**Table 1.** Characteristics of included studies.

| Study | Study type | Country | Population | Intervention/ control | Number of participants (males) | Age <sup>†</sup> | Outcome measures extracted | Total length of intervention (average) | Personnel administering treatment | Time elapsed since arriving in country (months) | Number of traumatic events | Maximum follow-up period (months) |
| --- | --- | --- | --- | --- | --- | --- | --- | --- | --- | --- | --- | --- |
| de Graaff et al. (2023) | RCT | Netherlands | Syrian refugees | PM+/CAU | 206 (127) | 36.4 ± 11.7 (18-69) | Depression (HSCL-25), anxiety (HSCL-25), PTSD (PCL-5) | 5 weeks | Trained peer refugees | 42.22 (1-97) | 9.90 ± 5.52 (0-26) | 3 |
| Eskici et al. (2023) | RCT | Turkey | Female Syrian refugees | CA-CBT/TAU | 23 (0) | 35.1 ± 8.3 | Depression (HSCL-25), PTSD (HTQ) | 7 weeks | Trained facilitators from local community | NR | NR | - |
| Jordans et al. (2023) | RCT | Lebanon | Syrian refugee adolescents | EASE/ETAU | 198 (101) | 11.8 ± 1.3 (10-14) | Depression (PHQ-A), PTSD (CRIS-13) | 7 weeks | Trained facilitators from local community | NR | 6.9 ± 3.9 (0-17) | 12 |
| Acarturk et al. (2022a) | RCT | Turkey | Syrian refugees | gPM+/ECAU | 46 (15) | 38.0 ± 10.9 | Depression (HSCL-25), anxiety (HSCL-25), PTSD (PCL-5) | 5 weeks | Trained peer refugees | NR | NR | 3 |
| Acarturk et al. (2022b) | RCT | Turkey | Syrian refugees | SH+/ECAU | 322 (116) | 31.2 ± 8.9 | Depression (PHQ-9), PTSD (PCL-5) | 5 weeks | Trained peer refugees | 54.53 ± 41.44 | NR | 6 |
| Bryant et al. (2022a) | RCT | Jordan | Syrian refugees | gPM+/EUC | 410 (110) | 40.0 ± 7.0 | Depression (HSCL-25), anxiety (HSCL-25), PTSD (PCL-5) | 5 weeks | Trained peer refugees | 70.7 ± 20 (12-108) | NR | 12 |
| Bryant et al. (2022b) | RCT | Jordan | Adolescent Syrian refugees | EASE/EUC | 471 (238) | 11.6 ± 1.3 (10-14) | Depression (PHQ-A), PTSD (CRIS-13) | 7 weeks | Trained facilitators from local community | NR | 6.9 ± 3.8 | 12 |
| Knefel et al. (2022) | RCT | Austria | Afghan refugees | aPM+/TAU | 51 (26) | 34.3 ± 13.6 | Depression (GHQ), anxiety (GHQ), PTSD (ITQ) | 6 weeks | Clinical psychologists | NR | 12.7 ± 5.4 | - |
| Orang et al. (2022) | RCT | Germany | Refugees of various nationalities | VBC/WLC | 103 (63) | 30.6 ± 8.1 (18-62) | Depression (PHQ-9), anxiety (GAD-7), PTSD (PCL-5) | 4 weeks (2-7 weeks) | Trained migrant counsellors (mostly refugees) | 34.3 ± 20.5 (1-480) | NR | 3 |
| Spaaij et al. (2022) | Pilot RCT | Switzerland | Syrian refugees | PM+/ETAU | 59 (29) | 39.9 ± 9.9 | Depression (HSCL-25), anxiety (HSCL-25), PTSD (PCL-5) | 5 weeks | Trained peer refugees | 40.3 ± 25.6 | 10.1 ± 5.03 | 3 |
| Aizik-Reebs et al. (2021) | RCT | Middle East (Israel) | Eritrean asylum seekers | MBTR-R/WLC | 158 (85) | 31.8 ± 5.21 (20-48) | Depression (PHQ-9), anxiety (Beck Anxiety Inventory), PTSD (HTQ) | 9 weeks | Clinical psychologists | NR | 6.16 ± 4.28 | 1.25 |
| Fine et al. (2021) | Cluster RCT | Tanzania | Adolescent Burundian refugees | EASE/ETAU | 82 (37) | 12.3 ± 1.5 (10-14) | PTSD (CPSS) | 7 weeks | Trained peer adult refugees | NR | NR | - |
| Foka et al. (2021) | CCT | Greece | Child refugees of various nationalities | SFJ/WLC | 72 (25) | 10.8 ± 2.0 (7-14) | Depression (CES-DC) | 6 days | Trained facilitator | NR | NR | - |
| Goninon et al. (2021) | CCT | Uganda | Congolese refugees | EMPOWER | 98 (45) | 33.4 ± 11.7 (18-80) | PTSD (SPTSS) | 2 weeks | Trained facilitator | 20.4 ± 24 | 8.45 ± 3.11 | 1.5 |
| Purgato et al. (2021) | RCT | Western European countries | Refugees of various nationalities | SH+/ETAU | 459 (325) | 32.6 ± 10.2 | Depression (PHQ-9), PTSD (PCL-5) | 5 weeks | Trained peer refugees | NR | NR | 12 |
| Alsheikh et al. (2020) | RCT | Jordan | Female Syrian refugees | Group counselling/ no counselling | 40 (0) | NR | PTSD (PCL-5) | 6 weeks | Clinical psychologist | NR | NR | - |
| Jeon et al. (2020) | CCT | South Korea | North Korean refugees | CBT/simple relaxation | 38 (3) | 38.1 (20-60) | Depression (CES-D), anxiety (STAI-S), PTSD (IES-R) | 8 weeks | Clinical psychologist and psychiatrist | NR | NR | - |
| Kananian et al. (2020) | RCT | Germany | Male Afghan and Iranian refugees | CA-CBT+/WLC/ECAU | 24 (24) | 21.9 ± 3.4 | PTSD (PCL-5) | 6 weeks | Trained facilitator | 21.8 ± 3.7 | NR | 12 |
| Tol et al. (2020) | Cluster RCT | Uganda | Female South Sudanese refugees | SH+/ECAU | 694 (0) | 30.9 ± 10.9 | Depression (PHQ-9), PTSD (PCL-C) | 5 weeks | Trained facilitators | NR | NR | 3 |
| Shaw et al. (2019) | RCT | Malaysia | Female Afghan refugees | CA-CBT/WLC | 29 (0) | 31.9 ± 9.9 | Depression (HSCL-25), anxiety (HSCL-25), PTSD (HTQ) | 8 weeks | Trained facilitators from local community | 21.6 | NR | 3 |
| Pfeiffer et al. (2018) | RCT | Germany | Child and adolescent refugees of various nationalities | Mein Weg/UC | 99 (92) | 17 ± 0.95 | Depression (PHQ-8), PTSD (CATS-S) | 6 weeks | Trained social workers | 12.7 ± 4.54 | NR | - |
| Ooi et al. (2016) | Cluster RCT | Australia | Child and adolescent refugees of various nationalities | TRT/WLC | 82 (29) | 12.6 ± 1.7 (10-17) | Depression (DSRS), PTSD (CRIS-13) | 8 weeks | Trained graduate psychology students | 28.1 ± 21.4 | 4.2 ± 2.1 (0-9) | 3 |
| Bolton et al. (2014) | RCT | Thailand | Burmese refugees | CETA/WLC | 347 (130) | 35.5 ± 12.1 (18-85) | Depression (HSCL-25), anxiety (HSCL-25), PTSD (HTQ) | 10 weeks (7-12 weeks) | Trained peer refugees | 66.5 ± 57.2 (0-420) | 12 ± 9 (1-24) | 4 |
| Hijazi et al. (2014) | RCT | USA | Iraqi refugees | Brief NET/WLC | 63 (28) | 48.2 ± 8.9 | Depression (Beck-II), PTSD (HTQ) | 3 weeks | Doctoral clinical psychology students | 27.6 ± 27.6 | 19.8 ± 6.4 | 4 |
| Stenmark et al. (2013) | RCT | Norway | Refugees of various nationalities | NET/TAU | 81 (56) | 35.3 ± 11.0 | Depression (HAM-D), PTSD (CAPS) | 10 weeks | Mental health professionals | 56 ± 50.6 | 8.2 ± 2.5 | 6 |
| Ertl et al. (2011) | RCT | Uganda | Former Ugandan child soldiers in IDP camps | NET/WLC | 57 (29) | 18.4 ± 3.6 | Depression (MINI), PTSD (CAPS) | 3 weeks | Trained facilitators from local community | - | 21.7 ± 4.2 | 12 |
| Sonderegger et al. (2011) | CCT | Uganda | Ugandan IDPs | EMPOWER/WLC | 202 (82) | 29.1 ± 12.6 | Depression (APAI *Two Tam, Kumu, Par*), anxiety (APAI *Ma Lwor*) | 2 weeks | Trained facilitators from local community | NR | NR | 3 |
| Neuner et al. (2010) | RCT | Germany | Refugees of various nationalities | NET/TAU | 32 (22) | 31.4 ± 7.6 | Depression (HSCL-25), PTSD (PDS) | 4 weeks (2.5-8.5 weeks) | Clinical psychologists | 55.6 ± 42.7 | NR | 6 |
| Ruf et al. (2010) | RCT | Germany | Child refugees of various nationalities | KIDNET/WLC | 26 (14) | 11.5 ± 3.1 (7-16) | PTSD (UPID) | 8 weeks | Clinical psychologists | 37.3 ± 46.9 | 4.4 ± 2.4 | 12 |
| Catani et al. (2009) | RCT | Sri Lanka | Child IDPs | KIDNET/MED-RELAX | 31 (17) | 12 ± 2 (8-14) | PTSD (UPID) | 2 weeks | Local counselor | NR | 4.6 ± 2 | 6 |
| Neuner et al. (2008) | RCT | Uganda | Rwandan and Somali refugees | NET/MG | 166 (83) | 34.8 ± 12.8 | PTSD (PDS) | 3 weeks | Trained lay counsellors | NR | 14 ± 4.8 | 8 |
| Ehntolt et al. (2005) | CCT | United Kingdom | Refugees of various nationalities | CBT/Control | 26 (17) | 14 ± 4.8 (11-15) | Depression (DSRS), anxiety (RCMAS), PTSD (R-IES) | 6 weeks | Clinical psychology trainee | NR | NR | - |
| Hinton et al. (2005) | RCT | USA | Cambodian refugees | CA-CBT/Delayed treatment | 40 (16) | 51.8 ± 6.8 | Anxiety (ASI), PTSD (CAPS) | 12 weeks | Psychiatrist | 206.8 ± 42.4 | NR | 6 |
| Hinton et al. (2004) | RCT | USA | Vietnamese refugees | CA-CBT/Delayed treatment | 12 (6) | NR | Depression (HSCL-25), anxiety (ASI), PTSD (HTQ) | 11 weeks | Psychiatrist | NR | NR | 9 |
| Neuner et al. (2004) | RCT | Uganda | Sudanese refugees | NET/SC | 31 (14) | 32.9 ± 6.8 | PTSD (PDS) | 2 weeks | Clinical psychologists | 39.4 ± 9.1 | 9.4 ± 6.6 | 12 |
| Barrett et al.<br>(2000) | CCT | Australia | Female teenage<br>refugees from<br>former<br>Yugoslavia | FRIENDS/<br>WLC | 20 (0) | 16.3 | Anxiety (SCAS) | 10 weeks | Clinical psychologist | 2.4 | NR | - |

<sup>†</sup> Ages are reported as mean ± SD, mean (range) or mean alone.
RCT, randomized controlled trial; CCT, controlled clinical trial.
IDP, internally displaced persons.
PM+, Problem Management Plus; gPM+, group PM+; CA-CBT, Culturally-Adapted Cognitive Behavioral Therapy; EASE, Early Adolescent Skills for Emotions; MBTR-R, Mindfulness-Based Trauma Recovery for Refugees; TRT, Teaching Recovery Techniques; KIDNET, Narrative Exposure Therapy for Traumatized Children and Adolescents
CAU, care as usual; TAU, treatment as usual; ETAU, enhanced treatment as usual; EUC, Enhanced Usual Care; MG, No-Treatment Monitoring Group; SC, Supportive Counseling.

**Table 2.** Reported outcomes of included studies.

| Study | Assessment measure used | Arm | No. of participants | Pre-intervention baseline | Post-intervention | 3-month follow-up | 6-month follow-up | 12-month follow-up |
| --- | --- | --- | --- | --- | --- | --- | --- | --- |
| de Graaff et al., 2023 | Anxiety (HSCL-25) | PM+ | 103 | 2.16 (0.66) | 1.85 (0.65) | 1.84 (0.64) | - | - |
|  |  | CAU | 103 | 2.24 (0.61) | 2.21 (0.64) | 2.15 (0.64) | - | - |
|  | Depression (HSCL-25) | PM+ | 103 | 2.39 (0.69) | 1.92 (0.64) | 1.91 (0.63) | - | - |
|  |  | CAU | 103 | 2.47 (0.72) | 2.33 (0.77) | 2.28 (0.69) | - | - |
|  | PTSD (PCL-5) | PM+ | 103 | 33.22 (17.84) | 20.89 (17.49) | 19.79 (16.59) | - | - |
|  |  | CAU | 103 | 35.57 (15.96) | 28.76 (16.52) | 28.21 (16.38) | - | - |
| Eskici et al., 2023 | Anxiety (HSCL-25) | CA-CBT | 12 | 2.39 (0.49) | 2.09 (0.51) | - | - | - |
|  |  | TAU | 11 | 2.48 (0.65) | 2.39 (0.70) | - | - | - |
|  | Depression (HSCL-25) | CA-CBT | 12 | 2.39 (0.49) | 2.09 (0.51) | - | - | - |
|  |  | TAU | 11 | 2.48 (0.65) | 2.39 (0.70) | - | - | - |
|  | PTSD (HTQ) | CA-CBT | 12 | 2.23 (0.43) | - | - | - | - |
|  |  | TAU | 11 | 2.1 (0.61) | - | - | - | - |
| Jordans et al., 2023 | Depression (PHQ-A) | EASE | 80 | 9.5 (0.8) | 8.3 (0.8) | 7.4 (0.9) | - | 9.7 (0.8) |
|  |  | ETAU | 118 | 9.6 (1.4) | 8.3 (1.4) | 8.3 (1.4) | - | 10.7 (1.4) |
|  | PTSD (CRIES-13) | EASE | 80 | 26.1 (2.2) | 26.7 (2.2) | 24.9 (2.3) | - | 28.2 (2.4) |
|  |  | ETAU | 118 | 26.3 (4.0) | 24.8 (4.0) | 25.0 (4.1) | - | 27.5 (4.1) |
| Acarturk et al. (2022a) | Anxiety (HSCL-25) | gPM+ | 24 | 2.37 (0.58) | 2.01 (0.59) | - | - | - |
|  |  | ECAU | 22 | 2.31 (0.64) | 2.01 (0.43) | - | - | - |
|  | Depression (HSCL-25) | gPM+ | 24 | 2.37 (0.58) | 2.01 (0.59) | 2.07 (0.52) | - | - |
|  |  | ECAU | 22 | 2.31 (0.64) | 2.01 (0.43) | 2.14 (0.43) | - | - |
|  | PTSD (PCL-5) | gPM+ | 24 | 1.84 (0.88) | 1.27 (0.7) | 1.12 (0.85) | - | - |
|  |  | ECAU | 22 | 1.7 (0.86) | 1.59 (0.86) | 1.26 (0.70) | - | - |
| Acarturk et al. (2022b) | Depression (PHQ-9) | SH+ | 322 | 6.45 (4.70) | 5.24 (4.91) | - | 4.93 (5.05) | - |
|  |  | ECAU | 320 | 6.30 (4.73) | 5.32 (5.12) | - | 6.70 (5.46) | - |
|  | PTSD (PCL-5) | SH+ | 322 | 20.72 (14.90) | 16.82 (12.83) | - | 13.99 (11.45) | - |
|  |  | ECAU | 320 | 20.14 (14.28) | 14.81 (14.60) | - | 15.09 (12.86) | - |
| Bryant et al. (2022a) | Anxiety (HSCL-25) | gPM+ | 204 | 24.81 (0.43) | 20.39 (0.5) | 20.03 (0.50) | - | 19.34 (0.54) |
|  |  | EUC | 206 | 25 (0.43) | 21.93 (0.48) | 19.65 (0.48) | - | 17.85 (0.50) |
|  | Depression (HSCL-25) | gPM+ | 204 | 36.57 (0.63) | 28.98 (0.73) | 29.04 (0.72) | - | 28.26 (0.78) |
|  |  | EUC | 206 | 35.15 (0.63) | 32.44 (0.70) | 31.30 (0.72) | - | 25.85 (0.73) |
|  | PTSD (PCL-5) | gPM+ | 204 | 25.98 (1.02) | 16.12 (1.05) | 10.31 (1.02) | - | 2.60 (1.07) |
|  |  | EUC | 206 | 26.72 (1.02) | 17.58 (1.01) | 10.73 (0.97) | - | 3.29 (1.00) |
| Bryant et al. (2022b) | Depression (PHQ-A) | EASE | 185 | 15.06 (0.43) | 12.80 (0.36) | 12.42 (0.35) | - | - |
|  |  | EUC | 286 | 15.39 (0.27) | 12.37 (0.29) | 12.36 (0.28) | - | - |
|  | PTS-D (CRIES-13) | EASE | 185 | 24.19 (0.89) | 18.39 (0.63) | 18.79 (0.72) | - | - |
|  |  | EUC | 286 | 23.29 (0.71) | 18.23 (0.5) | 18.87 (0.58) | - | - |
| Knefel et al. (2022) | Anxiety (GHQ) | aPM+ | 26 | 1.89 (0.43) | 1.37 (0.66) | - | - | - |
|  |  | TAU | 25 | 1.77 (0.75) | 1.86 (0.81) | - | - | - |
|  | Depression (GHQ) | aPM+ | 26 | 1.31 (0.63) | 0.84 (0.56) | - | - | - |
|  |  | TAU | 25 | 1.49 (0.94) | 1.41 (0.81) | - | - | - |
|  | PTSD (ITQ) | aPM+ | 26 | 13.67 (4.22) | 10.96 (5.5) | - | - | - |
|  |  | TAU | 25 | 14.8 (4.44) | 13.96 (5.98) | - | - | - |
| Orang et al. (2022) | Anxiety (GAD-7) | VBC | 42 | 13.54 (5.1) | 3.9 (4.41) | 2.51 (2.6) | - | - |
|  |  | WLC | 43 | 13.32 (5.78) | 14.81 (5.19) | - | - | - |
|  | Depression (PHQ-9) | VBC | 42 | 15.14 (6.09) | 4.40 (4.6) | 3.8 (3.6) | - | - |
|  |  | WLC | 43 | 16.27 (6.56) | 17.39 (6.49) | - | - | - |
|  | PTSD (PCL-5) | VBC | 28 | 44.7 (14.46) | 13.82 (10.78) | 10.03 (7.77) | - | - |
|  |  | WLC | 34 | 44.55 (14.9) | 48.17 (17.99) | - | - | - |
| Spaaij et al. (2022) | Anxiety (GAD-7) | PM+ | 31 | 22.45 (8.48) | 21 (7.6) | 19.55 (7.2) | - | - |
|  |  | ETAU | 28 | 19 (7.14) | 17.03 (8.43) | 18.05 (5.86) | - | - |
|  | Depression (PHQ-9) | PM+ | 31 | 35.41 (11.09) | 31.09 (7.53) | 31.61 (9.73) | - | - |
|  |  | ETAU | 28 | 29.86 (11.26) | 25.69 (9.1) | 28.18 (9.04) | - | - |
|  | PTSD (PCL-5) | PM+ | 31 | 39.26 (18.53) | 26.39 (16.62) | 28.51 (18.91) | - | - |
|  |  | ETAU | 28 | 26.63 (17.92) | 17.44 (16.19) | 20.42 (17.17) | - | - |
| Aizik-Reebs et al. (2021) | Anxiety (Beck Anxiety Inventory) | MBTR-R | 95 | 32.68 (9.25) | 23.53 (14.15) | - | - | - |
|  |  | WLC | 58 | 31.63 (13.18) | 31.62 (14.33) | - | - | - |
|  | Depression (PHQ-9) | MBTR-R | 95 | 15.12 (4.08) | 11.41 (5.76) | - | - | - |
|  |  | WLC | 58 | 16.03 (4.46) | 15.27 (5.16) | - | - | - |
|  | PTSD (HTQ) | MBTR-R | 95 | 2.71 (0.45) | 2.10 (0.59) | - | - | - |
|  |  | WLC | 58 | 2.85 (0.45) | 2.63 (0.48) | - | - | - |
| Fine et al. (2021) | PTSD (CPSS) | EASE |  |  |  |  |  |  |
|  |  | ETAU |  |  |  |  |  |  |
| Foka et al. (2021) | Depression (CES-DC) | SFJ | 33 | 17.09 (7.06) | 4.4 (4.39) | - | - | - |
|  |  | WLC | 39 | 17.13 (6.86) | 19.04 (5.96) | - | - | - |
| Purgato et al. (2021) | Depression (PHQ-9) | SH+ | 230 | 8.48 (5.82) | 6.10 (5.41) | - | 6.06 (5.57) | 4.03 (5.42) |
|  |  | ETAU | 229 | 8.38 (5.55) | 7.60 (5.82) | - | 6.49 (5.75) | 5.81 (6.05) |
|  | PTSD (PCL-5) | SH+ | 230 | 24.69 (16.35) | 19.22 (15.82) | - | 17.06 (16.10) | 12.45 (15.30) |
|  |  | ETAU | 229 | 22.77 (16.24) | 20.93 (17.00) | - | 16.79 (15.30) | 14.61 (15.31) |
| Alsheikh et al. (2020) | PTSD (PCL-5) | Group counselling | 20 | 57.65 (11.19) | 41.55 (7.11) | - | - | - |
|  |  | No counselling | 20 | 59.45 (8.01) | 58 (8.16) | - | - | - |
| Jeon et al. (2020) | Depression (CES-D) | CBT | 15 | 22.33 (13.87) | 16.4 (9.12) | - | - | - |
|  |  | Simple relaxation | 23 | 20.39 (13) | 19.61 (14.95) | - | - | - |
| Tol et al. (2020) | Depression (PHQ-9) | SH+ | 331 | 15.1 (4.7) | 9.7 (5.4) | 9.5 (4.2) | - | - |
|  |  | ECAU | 363 | 15.1 (4.8) | 12.8 (5.3) | 10.8 (5.1) | - | - |
|  | PTSD (PCL-C) | SH+ | 331 | 22 (4.7) | 16.1 (5.5) | 16.1 (4.9) | - | - |
|  |  | ECAU | 363 | 21.8 (4.8) | 19.2 (5.5) | 17.7 (5.8) | - | - |
| Shaw et al. (2019) | Anxiety (HSCL-25) | CA-CBT | 20 | 2.76 (0.49) | 2.17 (0.1) | 2.10 (0.16) | - | - |
|  |  | WLC | 9 | 2.71 (0.43) | 2.86 (0.14) | - | - | - |
|  | Depression (HSCL-25) | CA-CBT | 20 | 2.79 (0.61) | 1.99 (0.15) | 1.92 (0.16) | - | - |
|  |  | WLC | 9 | 2.8 (0.76) | 3.08 (0.22) | - | - | - |
|  | PTSD (HTQ) | CA-CBT | 20 | 2.58 (0.57) | 2.34 (0.13) | 2.15 (0.17) | - | - |
|  |  | WLC | 9 | 2.49 (0.71) | 3.01 (0.19) | - | - | - |
| Pfeiffer et al. (2018) | Depression (PHQ-8) | Mein Weg | 50 | 11.52 (0.71) | 8.25 (0.75) | - | - | - |
|  |  | UC | 49 | 11.47 (0.71) | 11.76 (0.76) | - | - | - |
|  | PTSD (CATS-S) | Mein Weg | 50 | 29.97 (1.22) | 23.52 (1.77) | - | - | - |
|  |  | UC | 49 | 31.85 (1.23) | 30.27 (1.73) | - | - | - |
| Ooi et al. (2016) | Depression (DSRS) | TRT | 44 | 10.96 (5.26) | 8.68 (5.48) | 8.29 (4.46) | - | - |
|  |  | WLC | 38 | 9.17 (4.61) | 8.03 (5.13) | - | - | - |
|  | PTSD (CRIES-13) | TRT | 43 | 23.02 (10.51) | 15.88 (9.58) | 12.71 (10.24) | - | - |
|  |  | WLC | 37 | 17.92 (11.86) | 14.17 (11.06) | - | - | - |
| Hijazi et al. (2014) | Depression (Beck-II) | Brief NET | 41 | 33.91 (10.46) | 2 months: 27.45 (13.54) | 4 months: 25.08 (13.27) | - | - |
|  |  | WLC | 22 | 33.45 (11.45) | 2 months: 31.45 (12.08) | 4 months: 27.38 (10.85) | - | - |
|  | PTSD (HTQ) | Brief NET | 41 | 2.79 (0.49) | 2 months: 2.60 (0.66) | 4 months: 2.55 (0.66) | - | - |
|  |  | WLC | 22 | 2.76 (0.44) | 2 months: 2.76 (0.48) | 4 months: 2.65 (0.52) | - | - |
| Ertl et al. (2011) | Depression (MINI) | NET | 29 | 2.76 (2.87) | - | 3.96 (2.72) | 3.08 (2.95) | 1.8 (2.18) |
|  |  | WLC | 28 | 1.71 (1.98) | - | 3.68 (2.86) | 3.21 (3.01) | 1.75 (2.56) |
|  | PTSD (CAPS) | NET | 29 | 67.03 (14.74) | - | 46.73 (19.24) | 43.00 (21.49) | 32.44 (22.84) |
|  |  | WLC | 28 | 63.61 (16.42) | - | 52.93 (20.83) | 48.61 (23.74) | 44.29 (29.56) |
| Neuner et al. (2010) | Depression (HSCL-25) | NET | 16 | 3.0 (0.4) | - | - | 2.6 (0.6) | - |
|  |  | TAU | 16 | 3.0 (0.5) | - | - | 2.9 (0.5) | - |
|  | PTSD (PDS) | NET | 16 | 38.9 (6.4) | - | - | 26.0 (9.2) | - |
|  |  | TAU | 16 | 36.9 (8.0) | - | - | 34.1 (6.1) | - |
| Ruf et al. (2010) | PTSD (UPID) | KIDNET | 13 | 43.3 (12.3) | 22.3 (18.6) | - | 17.2 (14.6) | 18.8 (14.8) |
|  |  | WLC | 13 | 38.3 (8.6) | - | - | 33.8 (17.4) | - |
| Catani et al. (2009) | PTSD (UPID) | KIDNET | 16 | 37.94 (14.8) | 12.41 (14.15) | - | 12.3 (10.87) | - |
|  |  | MED-RELAX | 15 | 36.56 (14.9) | 12.59 (11.06) | - | 9.75 (8.63) | - |
| Neuner et al. (2008) | PTSD (PDS) | NET | 111 | 25.9 (13.2) | - | 5.4 (6.6) | 6.1 (6.8) | - |
|  |  | MG | 111 | 26.7 (12.5) | - | 5.3 (5.7) | 5.0 (6.6) | - |
| Ehntolt et al. (2005) | Anxiety (RCMAS) | CBT | 15 | 16.87 (7.22) | 14.67 (7.12) | - | - | - |
|  |  | Control | 11 | 16.18 (6.57) | 8.91 (6.04) | - | - | - |
|  | Depression (DSRS) | CBT | 15 | 12.33 (4.7) | 11.67 (3.62) | - | - | - |
|  |  | Control | 11 | 12 (5.37) | 3 (6.57) | - | - | - |
|  | PTSD (R-IES) | CBT | 15 | 39.8 (8.4) | 33.8 (9.71) | - | - | - |
|  |  | Control | 11 | 38.55 (8.37) | 2.18 (9.38) | - | - | - |
| Hinton et al. (2005) | Anxiety (ASI) | CA-CBT | 20 | 3.08 (0.61) | 1.65 (0.45) | 1.86 (0.32) | 1.98 (0.40) | - |
|  |  | Delayed treatment | 20 | 3.27 (0.53) | 3.19 (0.36) | 1.91 (0.49) | - | - |
|  | PTSD (CAPS) | CA-CBT | 20 | 74.85 (14.67) | 39.25 (19.92) | 41.30 (13.95) | 44.59 (14.58) | - |
|  |  | Delayed treatment | 20 | 75.91 (11.5) | 73.05 (9.43) | 43.56 (10.22) | - | - |
| Hinton et al. (2004) | Anxiety (ASI) | CA-CBT | 6 | 43.5 (9.1) | 18.5 (6.4) | - | 8 months: 21.0 (8.0) | - |
|  |  | Delayed treatment | 6 | 38 (7.4) | 20.2 (4.9) | - | - | - |
|  | Depression (HSCL-25) | CA-CBT | 6 | 3.1 (0.7) | 2.1 (0.5) | - | 8 months: 1.6 (0.6) | - |
|  |  | Delayed treatment | 6 | 3.3 (0.9) | 3.2 (0.6) | - | - | - |
|  | PTSD (HTQ) | CA-CBT | 6 | 3.3 (0.4) | 1.7 (0.5) | - | 8 months: 1.8 (0.7) | - |
|  |  | Delayed treatment | 6 | 3.1 (0.7) | 3.3 (0.8) | - | - | - |
| Neuner et al. (2004) | PTSD (PDS) | NET | 17 | 25.2 (7.4) | 19.1 (11.7) | 4 months: 24.5 (7.8) | - | - |
|  |  | SC | 14 | 22.0 (8.0) | 19.8 (10.9) | 4 months: 22.8 (10.1) | - | - |
| Barrett et al. (2000) | Anxiety (SCAS) | FRIENDS | 9 | 39.89 (13.22) | 30.43 (11.37) | - | - | - |
|  |  | WLC | 11 | 30.64 (13.54) | 34.2 (8.45) | - | - | - |

Commonly used intervention programs included the WHO interventions Problem Management Plus (PM+; 5 studies) and Self-Help Plus (SH+; 3 studies) in adult samples; other interventions more generally employed the principles of Cognitive Behavioural Therapy (CBT; 7 studies) as well as Narrative Exposure Therapy (NET; 6 studies). In children/adolescents, Early Adolescent Skills for Emotions (EASE; 3 studies) and Narrative Exposure Therapy for Traumatised Children and Adolescents (KIDNET; 2 studies) were employed (see **Table 1**).

### Quality appraisal

Overall, 8 (27%) studies were reported as ‘low risk’, 16 (53%) had ‘some concern’, and 8 (27%) had ‘high risk’. The main areas of concern of the RCTs were missing outcome data and measurement of the outcome, where the psychological assessment tools used were not validated or even shown to be inconsistent. Analysis of the six remaining non-randomised studies using the ROBINS-I tool revealed methodological biases; four studies were reported as ‘serious risk’ and were judged ‘moderate risk’ (**SM 3**). The main flaws in the non-randomised trials were due to confounding, missing data, and measurement of outcomes.

The funnel plots of anxiety and PTSD outcomes were visually symmetrical (**SM 4**); Egger’s test for a regression intercept gave *p-*values of 0.63 and 0.07, respectively, indicating no evidence of publication bias. The funnel plot of depression outcomes showed asymmetry of effect sizes (Egger’s test *p* = 0.12), suggesting possible publication bias. A search of the WHO’s ICTRP (International Clinical Trials Registry Platform) did not show any unpublished studies.

### Effectiveness of psychosocial outcomes

**Figure 2** shows that brief psychological interventions were effective in decreasing anxiety symptoms (13 studies, 1183 participants) relative to controls (SMD −1.12, 95% CI −1.72 to −0.52, *I*^2^ = 95%). **Figure 3** shows that brief psychological interventions relative to control yielded standard mean differences −1.04, (95% CI −1.97 to −0.11, I^2^ 99%) for depression (20 studies, 3855 participants) and **Figure 4** shows the effectiveness for PTSD (24 studies, 3,770 participants), with a SMD of −0.82 (−1.20 to −0.45, I^2^ 93%).

**Figure 2.**
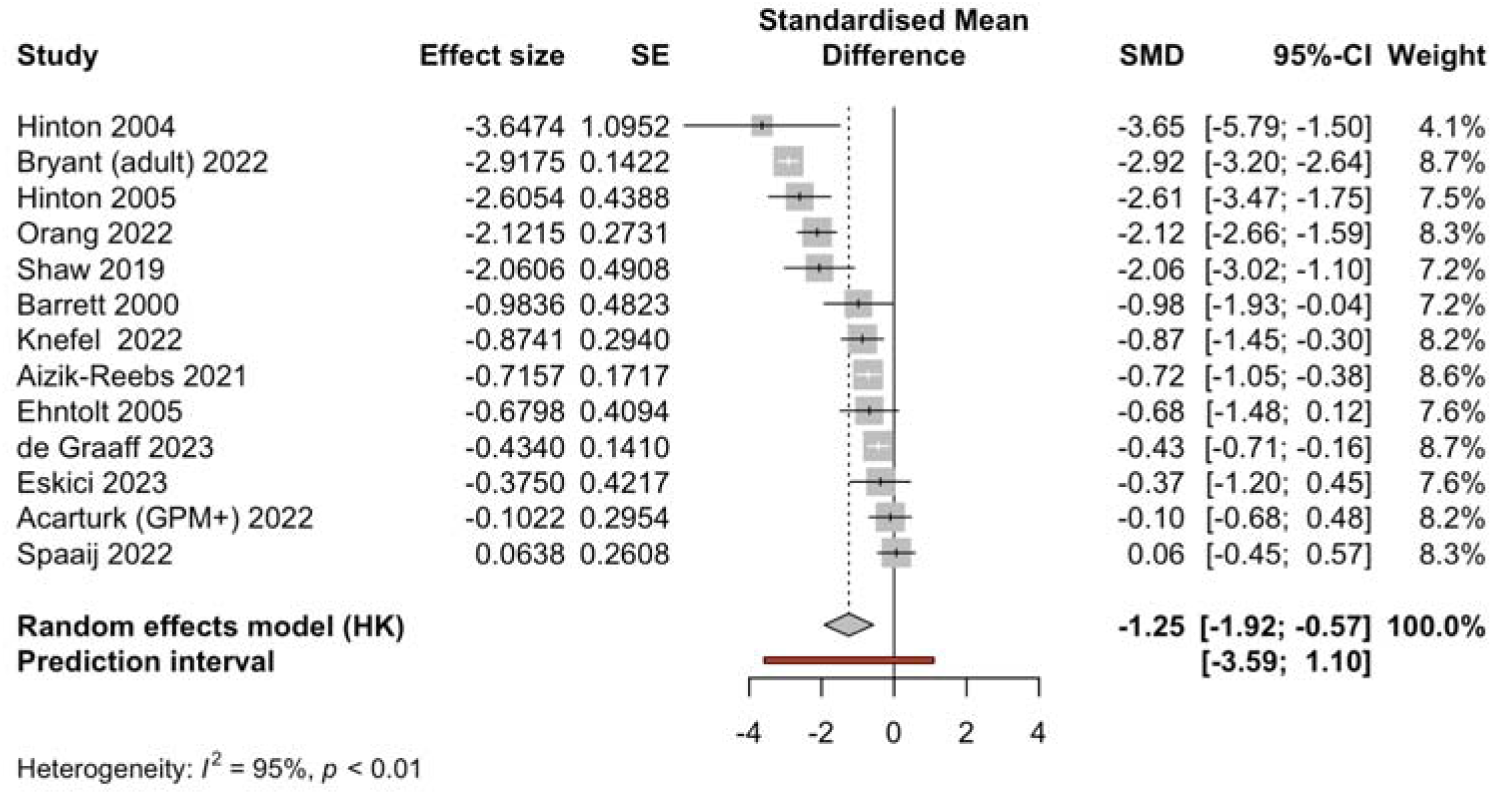
Effectiveness of brief CBT-based psychological interventions in treating anxiety in refugees and asylum seekers.

**Figure 3.**
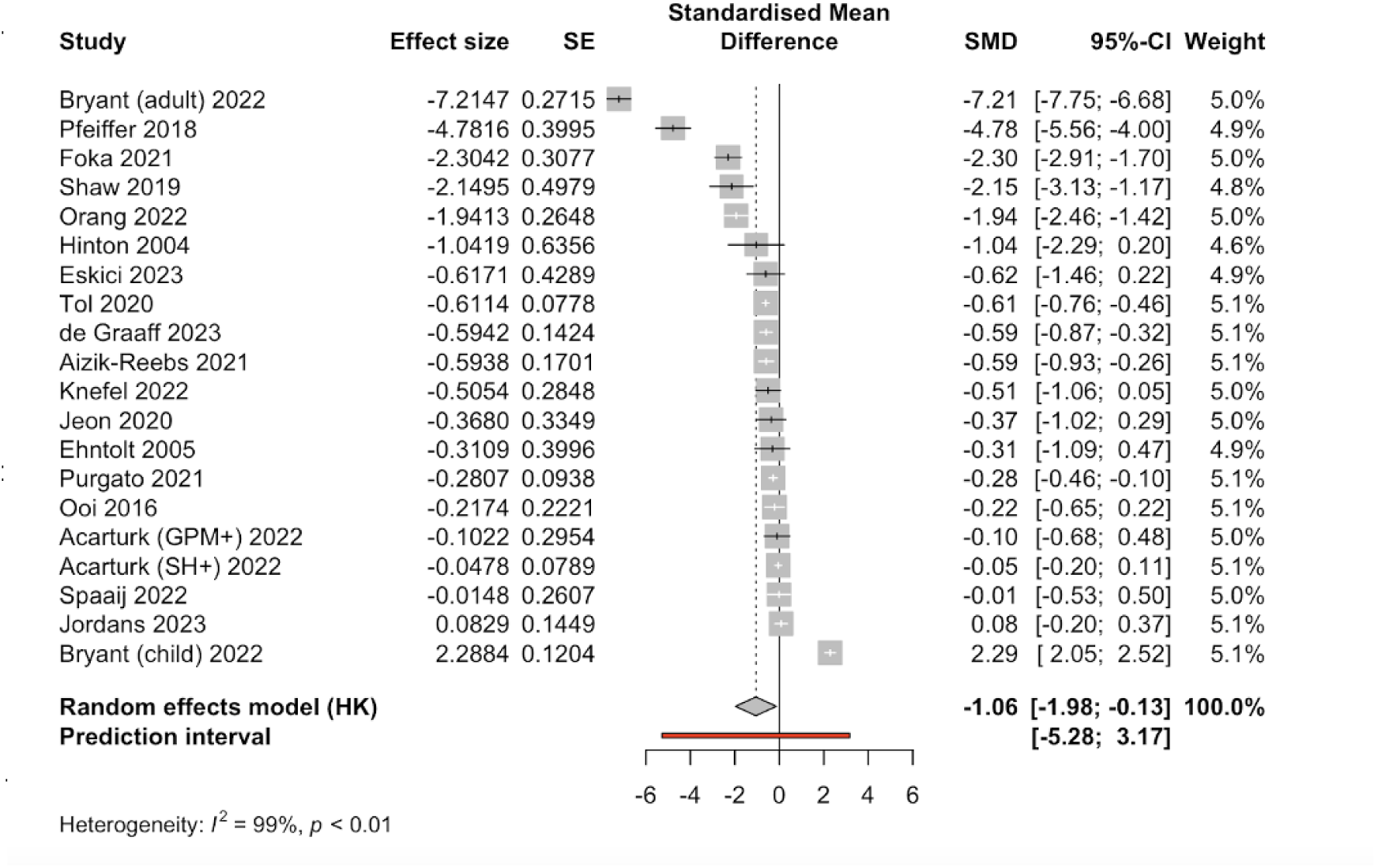
Effectiveness of brief CBT-based psychological interventions in treating depression in refugees and asylum seekers.

**Figure 4.**
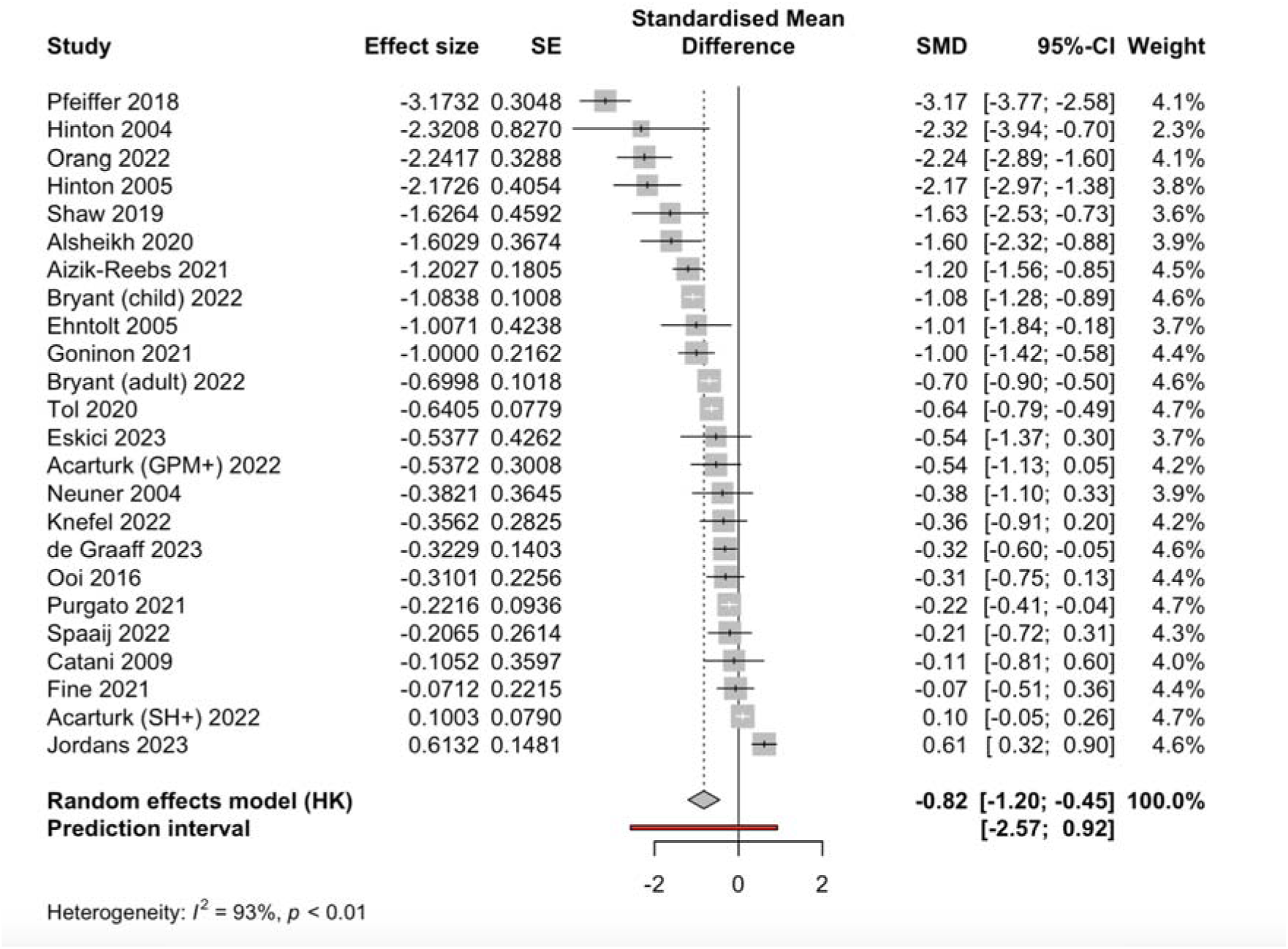
Effectiveness of brief CBT-based psychological interventions in treating PTSD in refugees and asylum seekers.

#### Intervention method

Four RCTs [Eskici et al., 2023; Shaw et al., 2009; Hinton et al., 2004, 2005] investigated the effectiveness of culturally adapted CBT (CA-CBT) sessions on anxiety; there was no significant effect (SMD −1.46, 95% CI −3.17 to 0.25, *I*^2^ = 83%). Three other RCTs (Spaaij et al., 2002; Knefel et al., 2022; de Graaff et al., 2023) utilised PM+ and did not demonstrate a significant effect (SMD −0.40, 95% CI −1.50 to 0.69, *I*^2^ = 66%). These same three studies using PM+ also found this intervention had no significant effect on depression (SMD −0.41, 95% CI −1.17 to 0.34, *I*^2^ = 48%). However, in the four RCTs [de Graaff et al., 2023; Spaaij et al., 2022; Knefel et al., 2022; Acarturk et al., 2022] using PM+, there was a significant effect on PTSD (SMD −0.34, 95% CI −0.50 to −0.17, *I*^2^ = 0%) (**SM 5.1**).

#### Intervention type (group vs individual)

We did not find any difference between interventions conducted in a group setting versus individually on anxiety, depression, and PTSD outcomes. (**SM 5.2**).

#### Intervention providers

We also examined the effectiveness of interventions conducted by trained lay persons (those in the local community or even peer refugees) versus professionals with a background in clinical psychology or psychiatry (**Figure 5**). Effect sizes were comparable for anxiety (lay persons: SMD −1.13, 95% CI −2.25 to −0.01, *I*^2^ = 97%; trained psychologists: SMD −1.09, 95% CI −1.85 to −0.33, *I*^2^ = 70%) and PTSD (lay persons: SMD −0.73, 95% CI −1.30 to −0.16, *I*^2^ = 95%; trained psychologists: SMD −0.95, −1.47 to −0.44, *I*^2^ = 76%). For depression, there was no significant difference between studies employing trained lay persons (SMD −1.27, 95% CI −2.63 to 0.08, *I*^2^ = 99%) compared to studies with interventions conducted by trained professionals (SMD −0.46, 95% CI −0.67 to −0.25, *I*^2^ = 0%).

**Figure 5.1.**
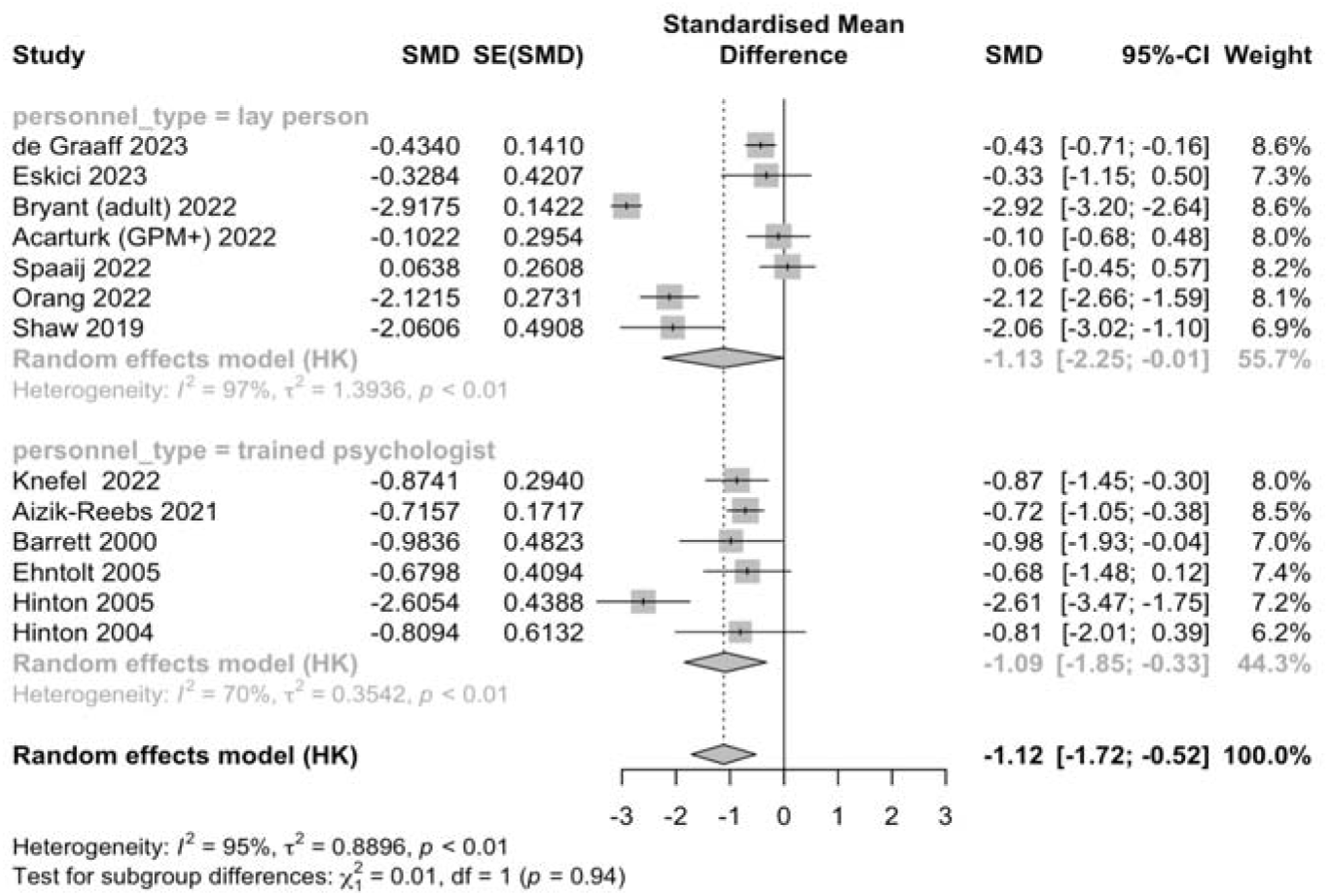
Effectiveness of interventions carried out by lay persons versus trained professionals in treating anxiety

**Figure 5.2.**
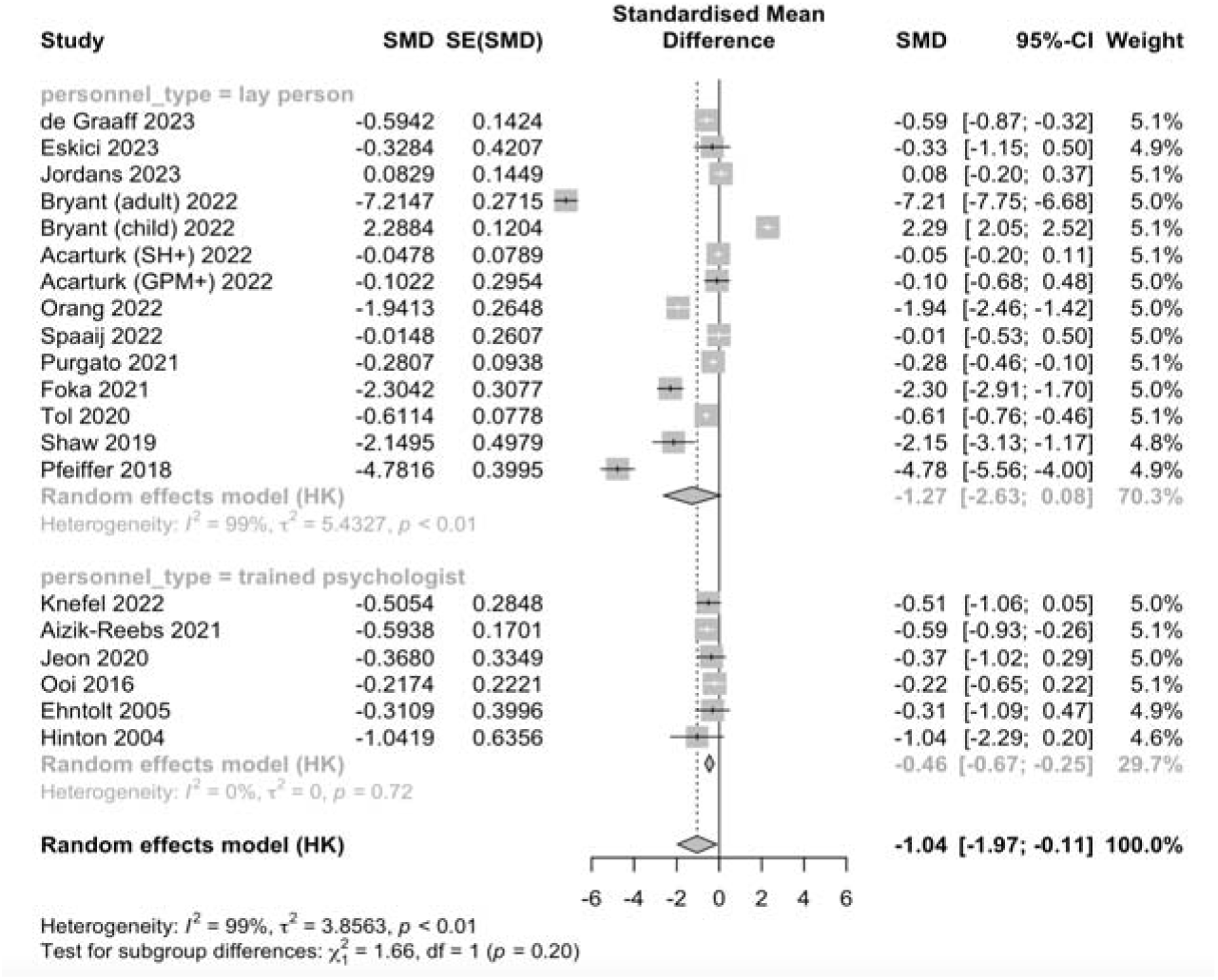
Effectiveness of interventions carried out by lay persons versus trained professionals in treating depression

**Figure 5.3.**
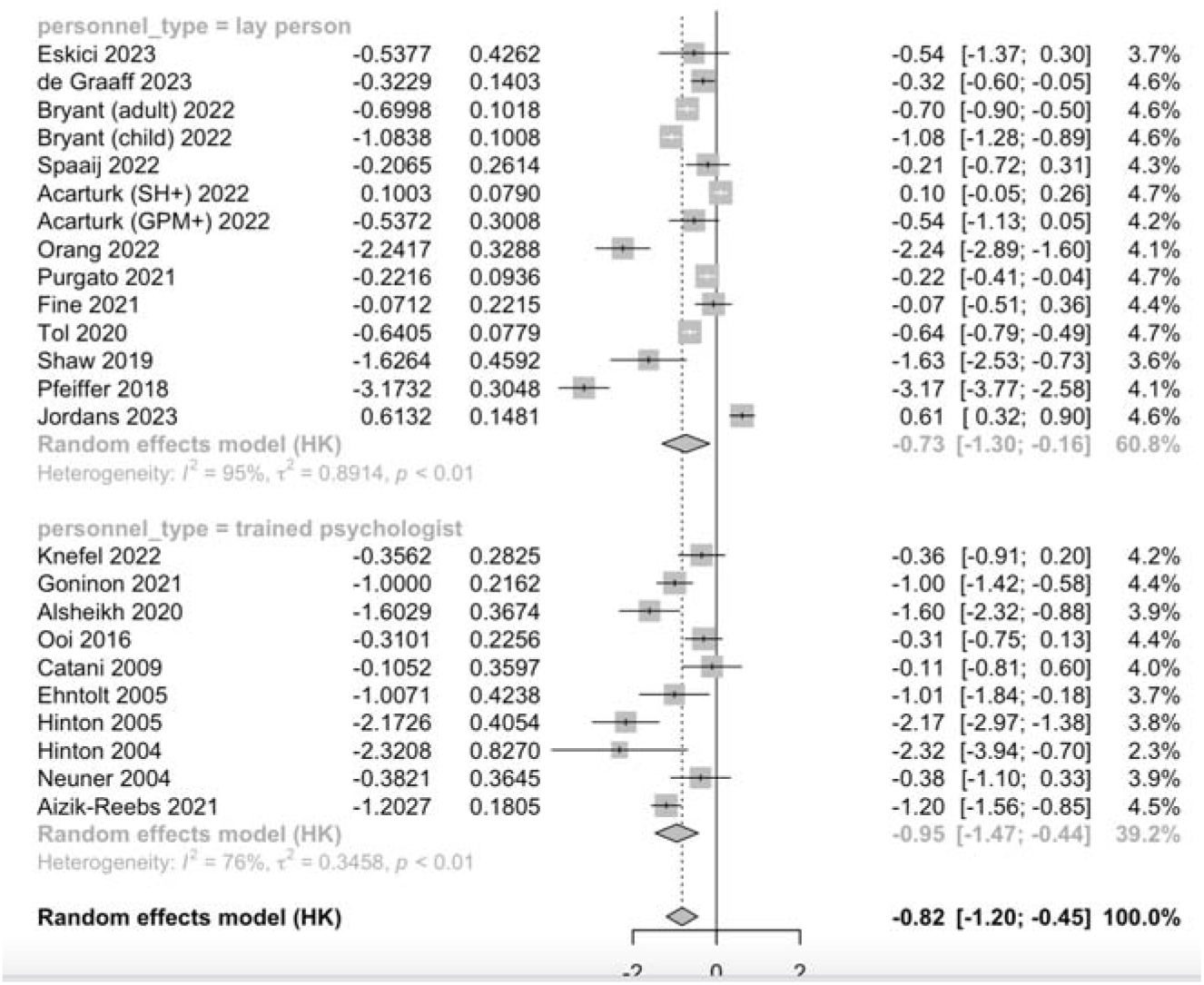
Effectiveness of interventions carried out by lay persons versus trained professionals in treating anxiety, depression, and PTSD

#### Intervention setting

We did not find any difference between interventions conducted in a low-to-medium resource setting versus a high-resource setting for anxiety, depression, and PTSD. (**SM 5.3**).

#### Population age

We divided the studies by whether they were performed on adults or children (8-18 years old) and found that positive effects of the intervention on anxiety were only present for adults (SMD −1.17, 95% CI −1.88 to −0.45, *I*^2^ = 96%), whereas children did not experience a significant change in their symptoms (SMD −0.81, 95% CI −2.71 to 1.10, *I*^2^ = 0%). This was also apparent for depression outcomes (adults: SMD −1.12, 95% CI −2.21 to −0.04, *I*^2^ = 98%; children: SMD −0.86, −3.38 to 1.67, *I*^2^ = 99%) and PTSD outcomes (adults: SMD −0.85, 95% CI −1.22 to −0.47, *I*^2^ = 90%; children: SMD −0.72, −1.85 to 0.41, *I*^2^ = 97%) (**Figure 6**).

**Figure 6.1.**
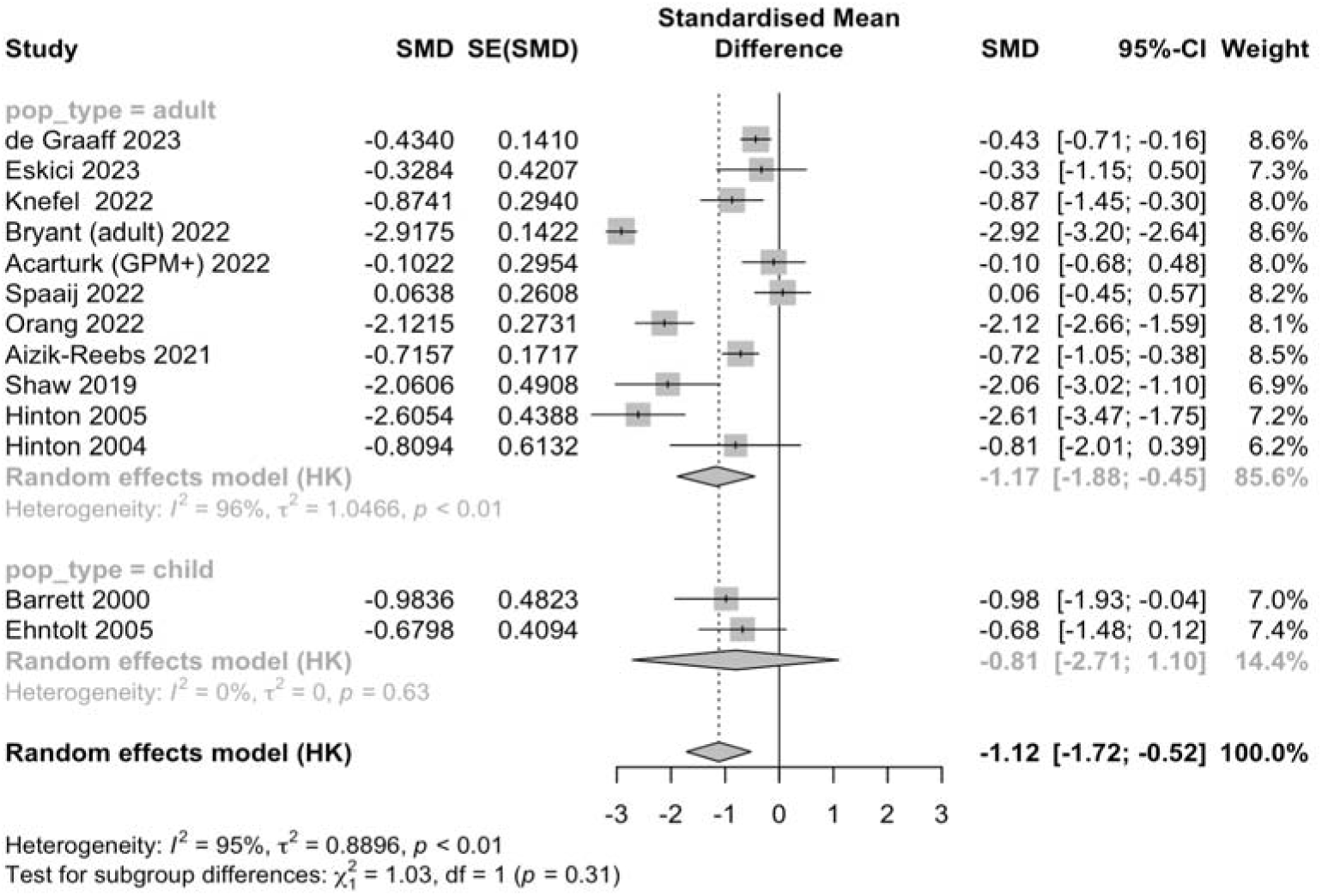
Effectiveness of interventions on adult and child populations for anxiety

**Figure 6.2.**
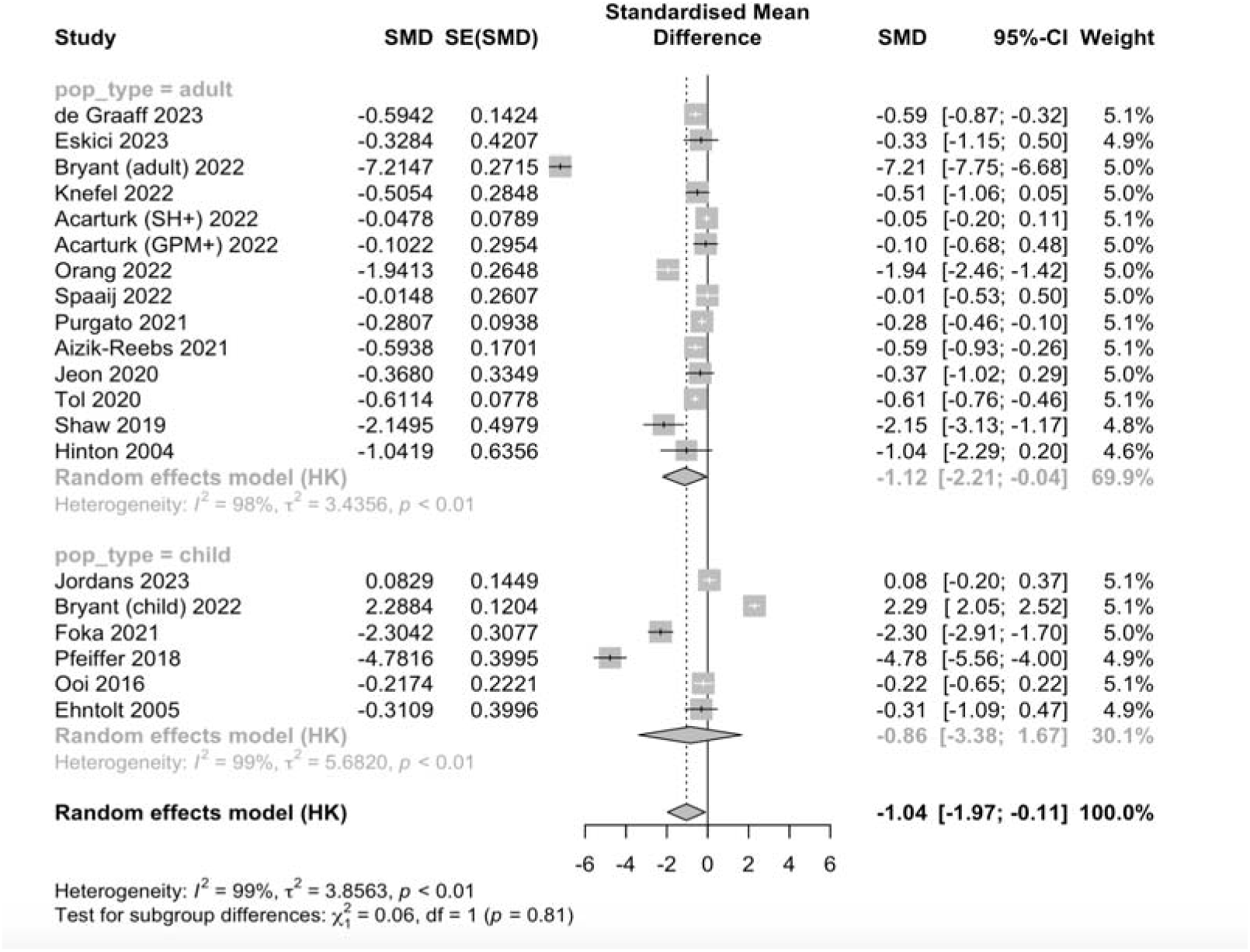
Effectiveness of interventions on adult and child populations for depression

**Figure 6.3.**
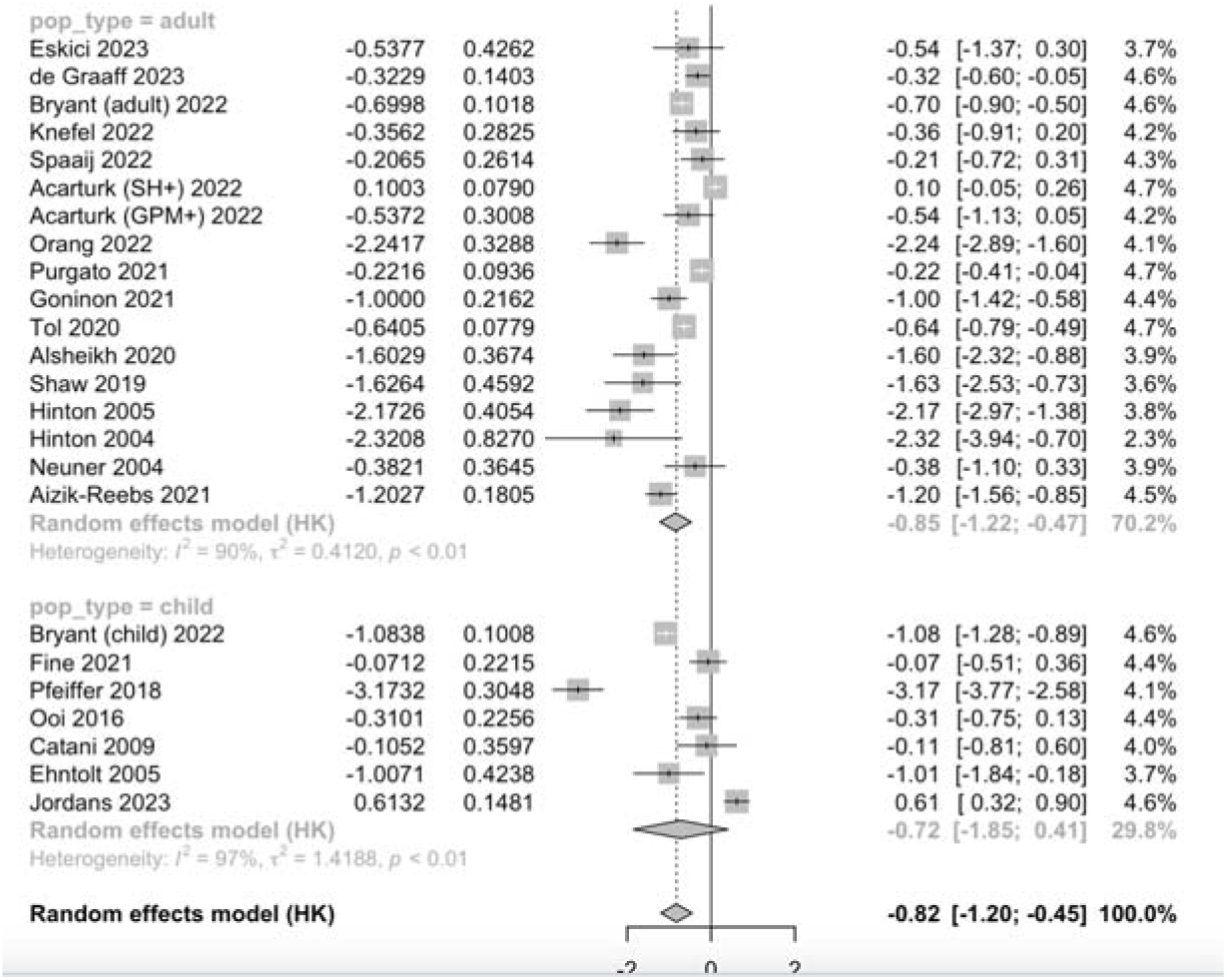
Effectiveness of interventions on adult and child populations for PTSD

#### Sample size

We did not find any difference between interventions conducted in a large patient population (*n* > 100) versus a smaller patient population (*n* < 100) on outcomes for anxiety, depression, and PTSD. (**SM 5.4**).

#### Type of mental health assessment tool

We did not find any difference between the type of mental health assessment tool that studies used to measure depression (e.g. PHQ-9), anxiety (e.g. HSCL-25), and PTSD outcomes (e.g. PCL-5). (**SM 5.5**). We did not show results on certain intervention methods (e.g. MBTR-R, Mein Weg) and types of mental health assessment tools (e.g. CES-D) as there were too few studies (< 3) to conduct a meta-analysis.

### Sensitivity analysis

Examining only studies that had low evidence of bias revealed a moderate positive effect of psychological interventions on PTSD (SMD −1.18, 95% CI −2.07 to −0.28, *I*^2^ = 86%) [Bryant et al., adult, 2022; Bryant et al., child, 2022; Tol et al., 2020; Hinton et al., 2004, 2005]. However, in the four studies assessing depression (Bryant et al., adult, 2022; Bryant et al., child, 2022; Tol et al., 2020; Hinton et al., 2004), we found no significant improvement in depression symptoms (SMD −1.64, 95% CI −8.02 to 4.73, *I*^2^ = 100%). Similarly, there was no significant effect of psychological interventions on anxiety (SMD −2.22, 95% CI −4.94 to 0.51, *I*^2^ = 82%) [Bryant et al., adult, 2022; Hinton et al., 2004, 2005] (**Figure 7**).

**Figure 7.1.**
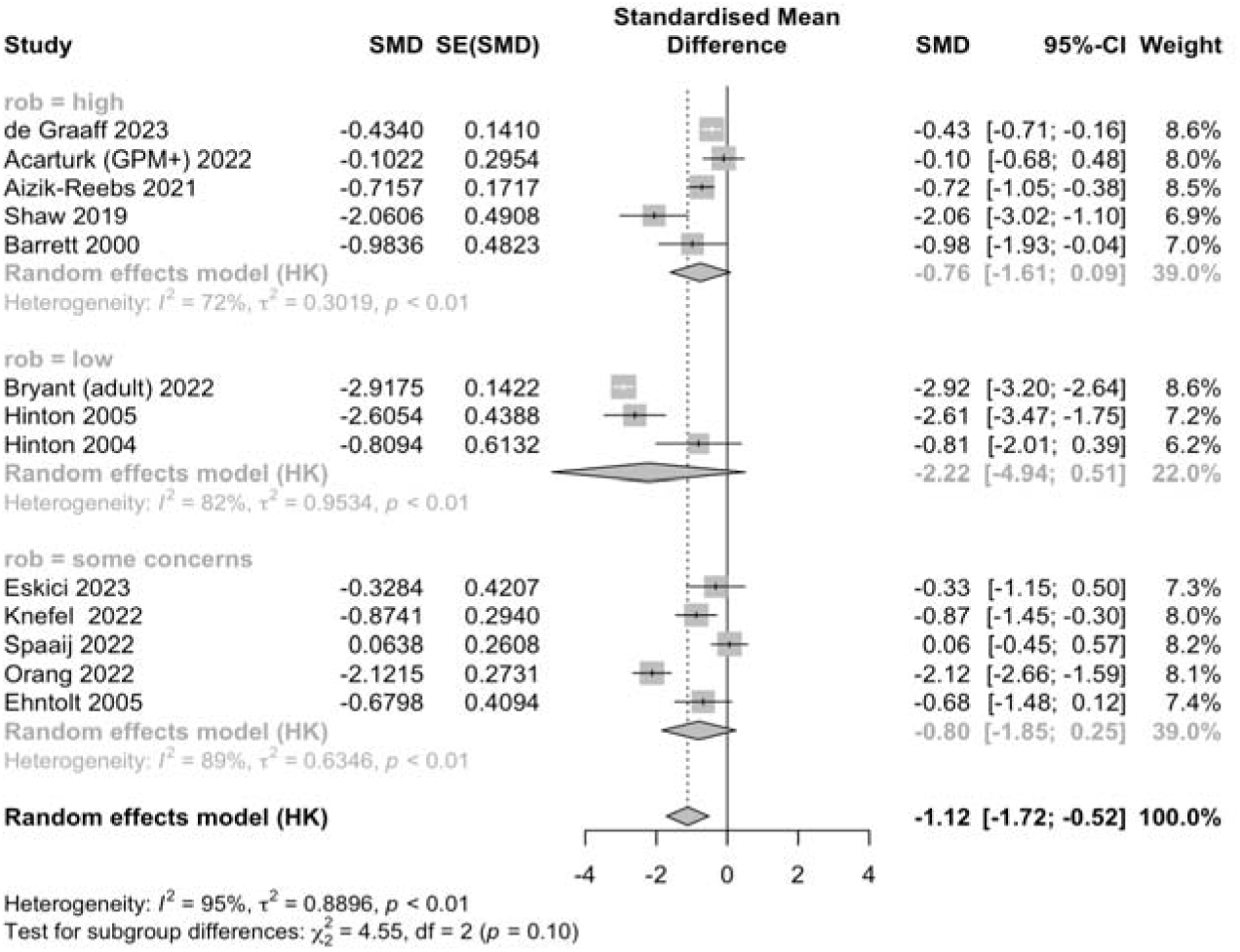
Sensitivity analysis: effectiveness of interventions carried out in studies with low, moderate, or high risk of bias for anxiety

**Figure 7.2.**
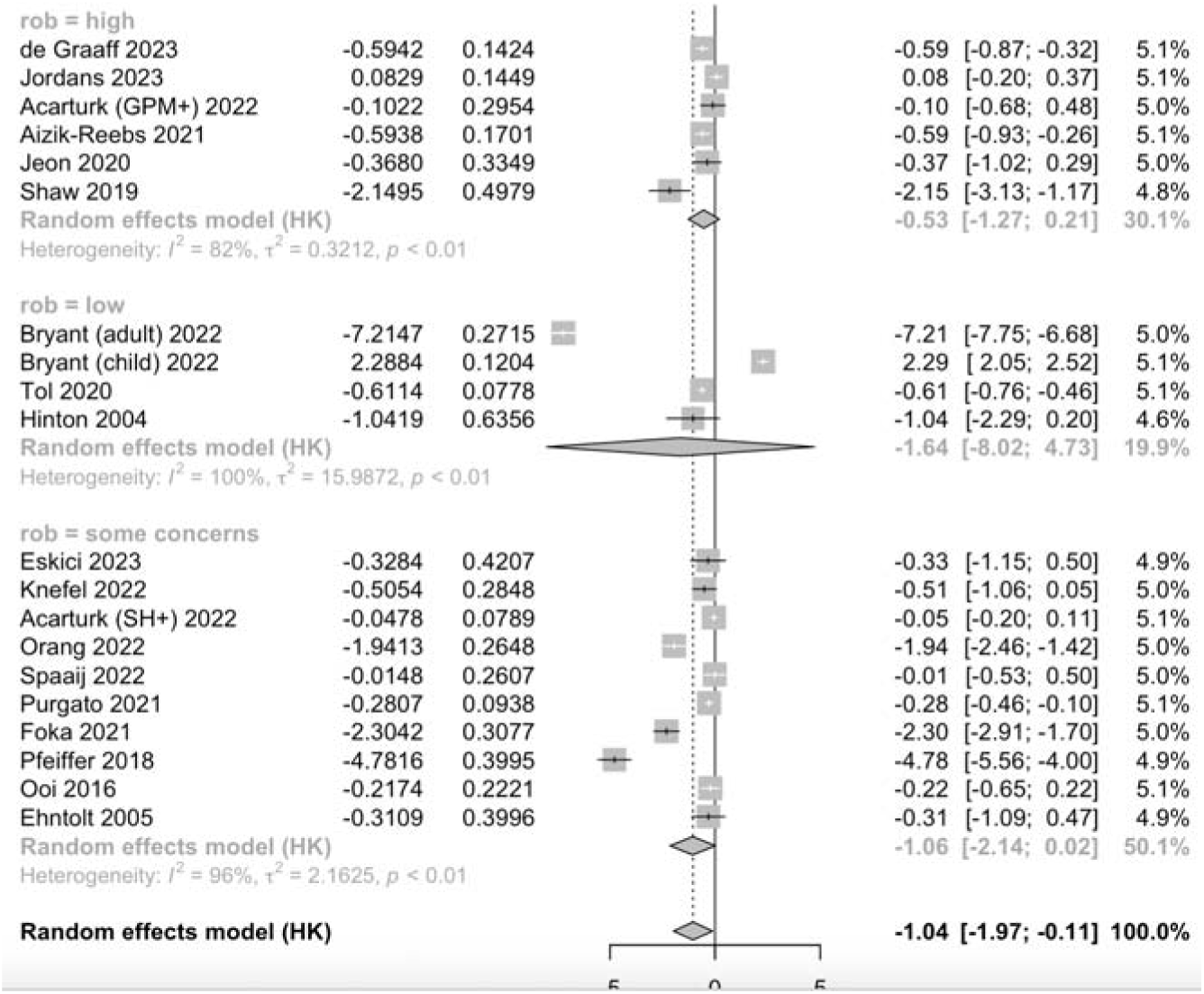
Sensitivity analysis: effectiveness of interventions carried out in studies with low, moderate, or high risk of bias for depression

**Figure 7.3.**
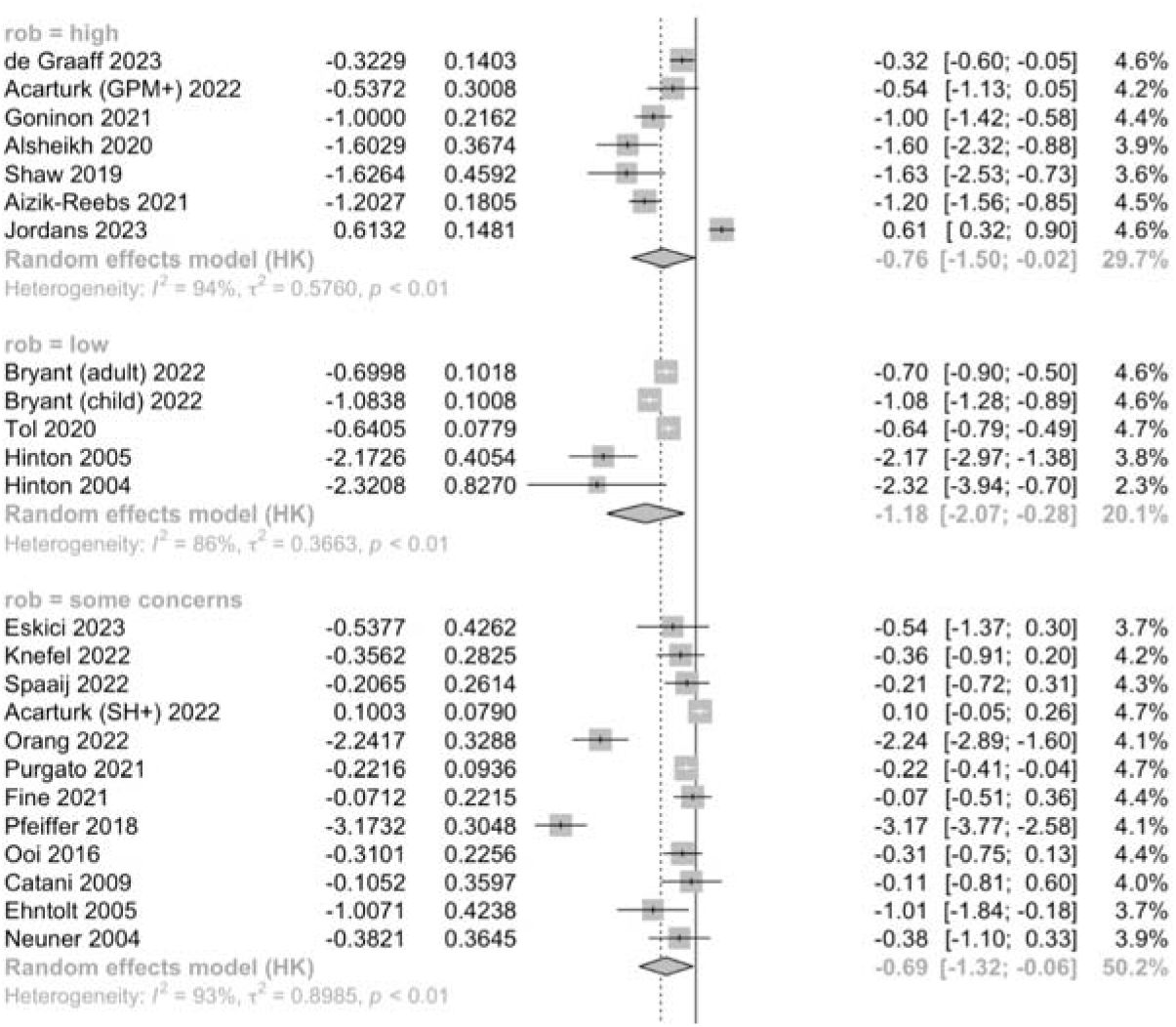
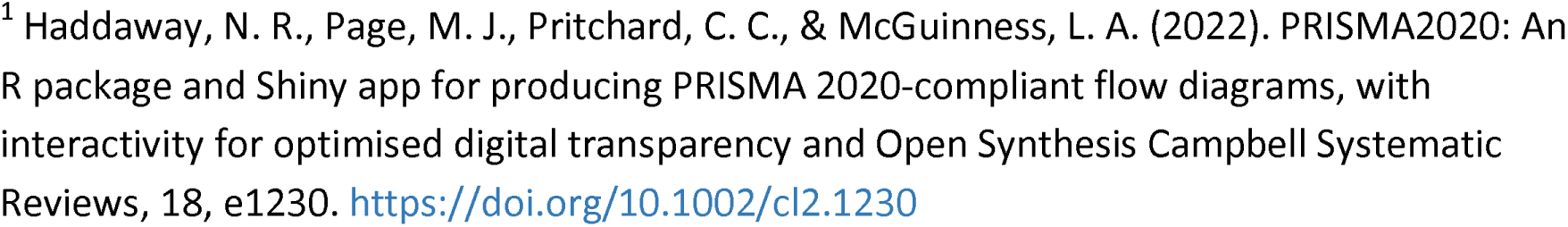
Sensitivity analysis: effectiveness of interventions carried out in studies with low, moderate, or high risk of bias for PTSD

### Long-term follow-up

Several studies reporting long-term follow-up were identified, and a post-hoc analysis of these outcomes was conducted. Follow-up durations were categorised into short-term (3-6 months) and long-term (7-12 months), in line with classifications used in similar systematic reviews ^21,31^. No study extended long-term follow-up beyond 12 months.

For short-term follow-up, there were four studies for anxiety, 9 for depression, and 12 for PTSD included in the analysis (**SM6**). The analysis revealed no significant effect for anxiety (SMD 0.24, 95% CI −0.94 to 1.42), depression (SMD −0.73, 95% CI −2.14 to 0.68), or PTSD (SMD 0.29, 95% CI −0.94 to 1.53).

For long-term follow-up, two studies reported results for depression, 1 for anxiety, and 3 for PTSD, with varying results. Bryant et al., 2022^32^ found no significant differences between treatment arms for depression, anxiety, and PTSD despite significant effects at post-intervention and short-term follow-up ^26^. Jordans et al., 2023^33^ reported no significant long-term effects for depression or PTSD. In contrast, Neuner et al., 2004^34^ found significant effects of the intervention on PTSD scores at 12 months.

## DISCUSSION

This systematic review aimed to investigate CBT-based psychological interventions conducted in a brief time period (under three months) in forcibly displaced persons. We found low-level evidence for the favourable effects of brief psychological interventions compared to (enhanced) care as usual on varied mental health outcomes. The results were unchanged even after removing low-quality studies for PTSD but not for depression and anxiety. The effects disappeared at short-term follow-up at 3-6 months and long-term follow-up 7-12 months. Subgroup analysis revealed a strong effect of brief interventions for adults compared with children and adolescents. Furthermore, there was no significant difference between interventions conducted by trained psychologists and those conducted by lay persons.

While the favourable effects of the brief CBT-based psychological interventions were found for PTSD, depression, and anxiety, the effects were greatest for anxiety compared to depression and PTSD, which suggests brief interventions have more of a role in the former. Studies were more likely to report PTSD compared to symptoms of depression and anxiety as compared to symptoms of PTSD. This may be due to the lower rates of anxiety and depression compared to PTSD found in the refugee population^7^. Previous reviews have found similar findings in refugee populations^11,21,22,24^; however, some only look at specific populations or single outcomes, or do not report the duration of interventional effects.

The interventional effects did not remain significant for anxiety, depression and PTSD in both short-term follow-ups of 3-6 months and in the majority of studies reporting long-term follow-ups of 7-12 months. This suggests that the benefits of these interventions may not endure despite the initial alleviation of symptoms, indicating that brief interventions are unable to comprehensively address the substantial mental health challenges faced by this population. The benefits on mental health symptoms may also diminish due to ongoing stressors experienced by forcibly displaced persons; for example, many studies took place in refugee camps, with well-documented stressors such as challenges meeting basic survival needs or unsafe living conditions^5,6,35^. Not all studies conducted short-term follow-ups, and only a few conducted long-term follow-ups. Additionally, there are often increasing losses to follow-up over time^32–34,36^. Therefore, more evidence is needed to investigate if the benefits of brief psychological interventions diminish over time. If confirmed, strategies should be identified to mitigate this decline, such as interventions administered over a longer timeframe or the provision of booster sessions.^37^

Analysis of subgroups comprising studies conducted by trained psychologists and briefly trained lay persons found no significant difference in the effects of these interventions. Furthermore, psychological interventions conducted by lay persons had significant beneficial effects on PTSD symptoms, which is consistent with previous systematic reviews ^21,22^. The effectiveness of these interventions may be attributed to the robustness of underlying therapeutic materials ^18,19^. The briefly trained lay persons in the studies were also often from the same background as those undergoing the intervention ^38–41^, thus, perhaps factors such as their relatability and cultural alignment with the community could contribute to the enhanced effectiveness of the interventions ^42^. Deploying briefly trained lay persons could reduce resource requirements, which is advantageous for the resource-limited settings where forcibly displaced persons are usually situated ^16,17^.

Subgroup analysis of age groups revealed no evidence for favourable effects of the brief CBT-based psychological interventions in children and adolescents compared with adults, in line with previous findings^22,43,44^. This is concerning given the high prevalence of mental health problems in forcibly displaced children and adolescents^9,45^, as well as evidence that evidence suggests that early interventions for children and adolescents can prevent long-term psychological problems^46,47^. The lack of significant effect in this demographic may stem from various factors. There was an overall poor quality of evidence, with the majority of studies involving this population (89%) receiving a risk of bias ratings of ‘some concerns’ or ‘high’. This may reflect the inherent challenges of conducting research involving children, such as ethical constraints related to utilisation of control groups for forcibly displaced youth. Additionally, the success of psychological interventions in children may be enhanced by parental involvement^46^, but this may be challenging in research due to the substantial number of unaccompanied and separated children in the refugee population^1,48^. Further research is critical to identify interventions that are effective for this age group and the determinants of their success.

### Limitations

The results of this study should be interpreted in light of some limitations. Firstly, our inclusion criteria were restricted to research papers published in English, and searches were conducted across only five databases, potentially resulting in the omission of relevant studies. While efforts were made to mitigate this through a comprehensive search strategy (e.g. extensive citation searching of related systematic reviews and included studies), it is possible that other relevant studies exist that were not captured by our search.

Our decision to impose a 3-month time limit for the duration of psychological intervention may have led to the exclusion of relevant studies. However, this limit was chosen to align with the focus on brief psychological interventions, which are commonly defined as interventions lasting less than 3 months. Brief psychological interventions may be more suitable for forcibly displaced persons, given the transient nature of their living situations and limited mental health resources available. An additional limitation arose from inconsistencies in the description of interventions and timing across many studies; therefore, we had to exclude studies which may have contained relevant evidence.

We found high statistical heterogeneity across all analyses. This could be attributed to multiple factors, including intervention type, intervention setting, and demographic characteristics of the study population. We performed sensitivity analysis by the quality of the study, which removed the heterogeneity for anxiety; however, only three studies were considered high quality. However, we reported the pooled estimates as the majority of studies lie in the same direction, including studies with non-significant results. When restricting this to studies with significant effects, i.e., not crossing the null line, 9/13 studies for anxiety, 9/20 studies for depression, and 14/24 studies for PTSD found a positive outcome.

Overall, the risk of bias was high, which undermined our confidence in the results. This may reflect challenges associated with conducting research with forcibly displaced populations, such as high dropout rates. Both randomised and non-randomised trials were included so as not to exclude relevant research where randomisation was not feasible due to lack of resources; however, this may have biased results. Finally, digital interventions, which may be more effective and less resource-intensive than face-to-face interventions, were excluded.^50^

### Implications for Practice, Policy, and Research

There is sufficient evidence to warrant brief psychological interventions for refugee populations. It is more likely that these interventions have an impact on anxiety and PTSD, whereas, without ongoing support, depression symptoms are unlikely to improve. Our findings suggest that to optimise the use of resources, brief psychological interventions can be delivered by lay people, and policymakers should consider how refugees themselves can be trained to deliver such interventions to improve the mental health outcomes of refugee populations.

From a research perspective, there is a need to undertake high-quality trials with long-term follow-up and develop reporting standards that facilitate improved descriptions of the interventions and core outcomes to aid pooled analysis. This is a particularly pressing issue in the forcibly displaced youth, in whom interventions have generally been found to be ineffective. The role of digital interventions warrants further research exploring their performance as well as accessibility and applicability.

## CONCLUSION

This systematic review and meta-analysis presented a comprehensive and robust synthesis of the use of brief psychological interventions to improve mental health outcomes in forcibly displaced people. The results of this study showed low-level evidence that brief psychological interventions have positive effects in forcibly displaced persons as compared to control. There was considerable heterogeneity among studies, which remained unexplained even after conducting several subgroup and sensitivity analyses. Subgroup analysis showed that while brief psychological interventions were effective in adults, they did not appear to be effective in children and adolescents. There was no significant difference observed between interventions conducted by mental health professionals and those by briefly trained peer refugees/lay persons. The effects of the interventions did not appear to translate into long-term positive effects on mental health at 3 months and beyond.

These findings point towards the feasibility of training peer refugees to deliver psychological interventions in lower-resource settings, a need for more rigorous studies in child and adolescent populations, and more sustained intervention in order to produce positive long-term psychological effects.

## Data Availability

All data produced in the present study are available upon reasonable request to the authors

## Acknowledgements

We are most grateful to Georgia Richards for her guidance with publishing the protocol and Nia Roberts for her assistance in drafting our initial search strategy. We would also like to thank all the authors of the primary studies who kindly provided additional information upon request.

## Author contributions

Conception and design: ND, LX, EED, ET, CH

Data collection: ND, LX, EED

Data analysis and interpretation: ND, LX, ET, EA

Writing (original draft): ND, LX

Review and editing: ND, LX, ET, CH

