## Supplementary Material for "Brief psychological interventions to improve mental health outcomes in refugee populations: A systematic review"

**Table of Contents**

SM1. Differences between protocol and review

SM2. Search strategies

SM3. Risk of bias

SM4. Funnel plots

SM5. Subgroup analysis

SM6. Long term analysis

**SM1. Differences between protocol and review**

(preregistration-ID: 10.17605/OSF.IO/9CXU4)

Analysis of primary outcomes revealed high heterogeneity in the results. Therefore, though this was not in our protocol, we performed subgroup analyses to investigate causes of heterogeneity, grouping studies by the type of mental health assessment tools used, intervention type, population type, personnel conducting the intervention, sample size (>100 or <100), and whether the setting was in a low or high-income country. We also performed sensitivity analysis by removing studies with a moderate and high risk of bias.

During data extraction, we found a number of studies which included long-term follow up. Therefore, though this was not in our protocol, we extracted and analysed long-term results.

**SM2. Search strategies**

**Ovid Medline, EMBASE**

1	(Refugee* or migrant* or immigrant* or "asylum seeker*").mp.

2	(displaced adj3 (population* or person* or group* or individual*)).mp.

3	(undocumented adj3 (population* or person* or group* or individual*)).mp.

4 (Counselling or Counseling or "cognitive behavioural therapy" or CBT).mp.

5	(Intervention? adj3 (psycholog* or psychiat* or behavior* or behaviour* or cognit*)).mp.

6	(Therap* adj3 (psycholog* or psychiat* or behavior* or behaviour* or cognit*)).mp. 7	(Program? adj3 (psycholog* or psychiat* or behavior* or behaviour* or cognit* or intervention? or therap*)).mp.

8	undocumented immigrants/ or refugees/

9	(Depressi* or anxi*).mp.

10	("Post traumatic stress disorder" or "post-traumatic stress disorder" or PTSD).mp.

11	(Disorder* adj3 (depressi* or anxi* or mood or psychiatr* or psychologic*)).mp.

12	(distress adj3 (psycholog* or psychiatr*)).mp.

13	exp community psychiatry/ or exp mental health services/ or community mental health services/ or exp counseling/ or exp emergency services, psychiatric/ or social work, psychiatric/ or exp psychotherapy/ or exp cognitive behavioral therapy/ or exp relaxation therapy/ or emotion-focused therapy/ or psychotherapy, brief/ or psychotherapy, multiple/

14	mental health/ or exp anxiety disorders/ or exp mood disorders/ or exp "bipolar and related disorders"/ or exp depressive disorder/ or exp "trauma and stressor related disorders"/ or adjustment disorders/ or exp stress disorders, traumatic/

15	1 or 2 or 3 or 8

16	4 or 5 or 6 or 7 or 13

17	9 or 10 or 11 or 12 or 14

18	15 and 16 and 17

**OVID PsycINFO**

1	(Refugee* or migrant* or immigrant* or "asylum seeker*").mp.

2	(displaced adj3 (population* or person* or group* or individual*)).mp.

3	(undocumented adj3 (population* or person* or group* or individual*)).mp.

4	exp refugees/

5	(Intervention? adj3 (psycholog* or psychiat* or behavior* or behaviour* or cognit*)).mp.

6	(Therap* adj3 (psycholog* or psychiat* or behavior* or behaviour* or cognit*)).mp.

7	(Program? adj3 (psycholog* or psychiat* or behavior* or behaviour* or cognit* or intervention? or therap*)).mp.

8	exp community mental health services/

9	exp cognitive behavior therapy/

10	exp trauma treatment/

11	(Depressi* or anxi*).mp.

12	("Post traumatic stress disorder" or "post-traumatic stress disorder" or PTSD).mp.

13	(Disorder* adj3 (depressi* or anxi* or mood or psychiatr* or psychologic*)).mp.

14	(distress adj3 (psycholog* or psychiatr*)).mp.

15	exp "stress and trauma related disorders"/ or exp acute stress disorder/ or exp adjustment disorders/ or exp attachment disorders/ or exp posttraumatic stress disorder/

16	exp anxiety disorders/

17	major depression/

18	exp "depression (emotion)"/

19	1 or 2 or 3 or 4

20	5 or mk6 or 7 or 8 or 9 or 10

21	11 or 12 or 13 or 14 or 15 or 16 or 17 or 18

22	19 and 20 and 21

23	22 and 2000:2023.(sa_year).

**CINAHL**

S1	(MH "Refugees") OR (MH "Transients and Migrants") OR (MH "Immigrants+") OR (MH "Refugee Camps")

S2	Refugee* OR migrant* OR immigrant* OR "asylum seeker*"

S3	displaced N3 (population* OR person* OR group* OR individual*)

S4	undocumented N3 (population* OR person* OR group* OR individual*)

S5	(MH "Undocumented Immigrants")

S6	S1 OR S2 OR S3 OR S4 OR S5

S7	(MH "Community Mental Health Services+") OR (MH "Social Work, Psychiatric") OR (MH "Psychological First Aid") OR (MH "Behavior Therapy+") OR (MH "Psychotherapy+") OR "Counselling OR Counseling OR "cognitive behavioural therapy" OR CBT"

S8	Counselling or Counseling or "cognitive behavioural therapy" or CBT OR Intervention# N3 (psycholog* OR psychiat* OR behavior* OR behaviour* OR cognit*)

S9	Therap* N3 (psycholog* OR psychiat* OR behavior* OR behaviour* OR cognit*)

S10	Program# N3 (psycholog* OR psychiat* OR behavior* OR behaviour* OR cognit* OR intervention# OR therap*)

S11	(MH "Stress Management")

S12	S7 OR S8 OR S9 OR S10 OR S11

S13	Depressi* OR anxi*

S14	(MH "Stress Disorders, Post-Traumatic+") OR (MH "Anxiety Disorders+") OR (MH "Psychological Trauma+") OR (MH "Adjustment Disorders+") OR (MH "Mental Disorders, Chronic") OR (MH "Affective Disorders, Psychotic+") OR (MH "Schizoaffective Disorder") OR (MH "Psychiatric Emergencies") OR (MH "Depression+") OR (MH "Dissociative Disorders+") OR (MH "Affective Symptoms+") OR (MH "Self-Injurious Behavior") OR (MH "Self Neglect") OR (MH "Stress, Psychological+") OR (MH "Suicidal Ideation") OR (MH "Suicide, Attempted") OR (MH "Schizophrenia+")

S15	"Post traumatic stress disorder" OR "post-traumatic stress disorder" OR PTSD OR “post traumatic stress syndrome” OR “post-traumatic stress syndrome” OR PTSS

S16	Disorder* N3 (depressi* OR anxi* OR mood OR psychiatr* OR psychologic*) OR distress adj3 (psycholog* or psychiatr*)

S17	S13 OR S14 OR S15 OR S16

S18	S6 AND S12 AND S17

**Global Index Medicus**

tw:((tw:(tw:((tw:(depressi* OR anxi*)) OR (tw:("Post traumatic stress disorder" OR "post-traumatic stress disorder" OR ptsd OR “post traumatic stress syndrome” OR “post-traumatic stress syndrome” OR ptss)) OR (tw:(disorder* adj3 (depressi* OR anxi* OR mood OR psychiatr* OR psychologic*))) OR (tw:(distress adj3 (psycholog* OR psychiatr*)))))) AND (tw:(tw:((tw:(tw:((tw:(refugee* OR migrant* OR immigrant* OR "asylum seeker*")) OR (tw:(displaced adj3 (population* OR person* OR group* OR individual*))) OR (tw:(undocumented adj3 (population* OR person* OR group* OR individual*)))))) OR (tw:(( mh:("Refugees"))))))) AND (tw:(tw:((tw:((tw:(counselling OR counseling OR "cognitive behavioural therapy" OR "cognitive behavioral therapy" OR cbt )) OR (tw:(intervention* adj3 (psycholog* OR psychiat* OR behavior* OR behaviour* OR cognit*))) OR (tw:(therap* adj3 (psycholog* OR psychiat* OR behavior* OR behaviour* OR cognit*))) OR (tw:(program* adj3 (psycholog* OR psychiat* OR behavior* OR behaviour* OR cognit* OR intervention* OR therap*))))) OR (tw:(( mh:("Mental Health Services" OR "Emergency Services, Psychiatric" OR "Social Work, Psychiatric" OR "counseling"))))))))

**SM3. Risk of bias**

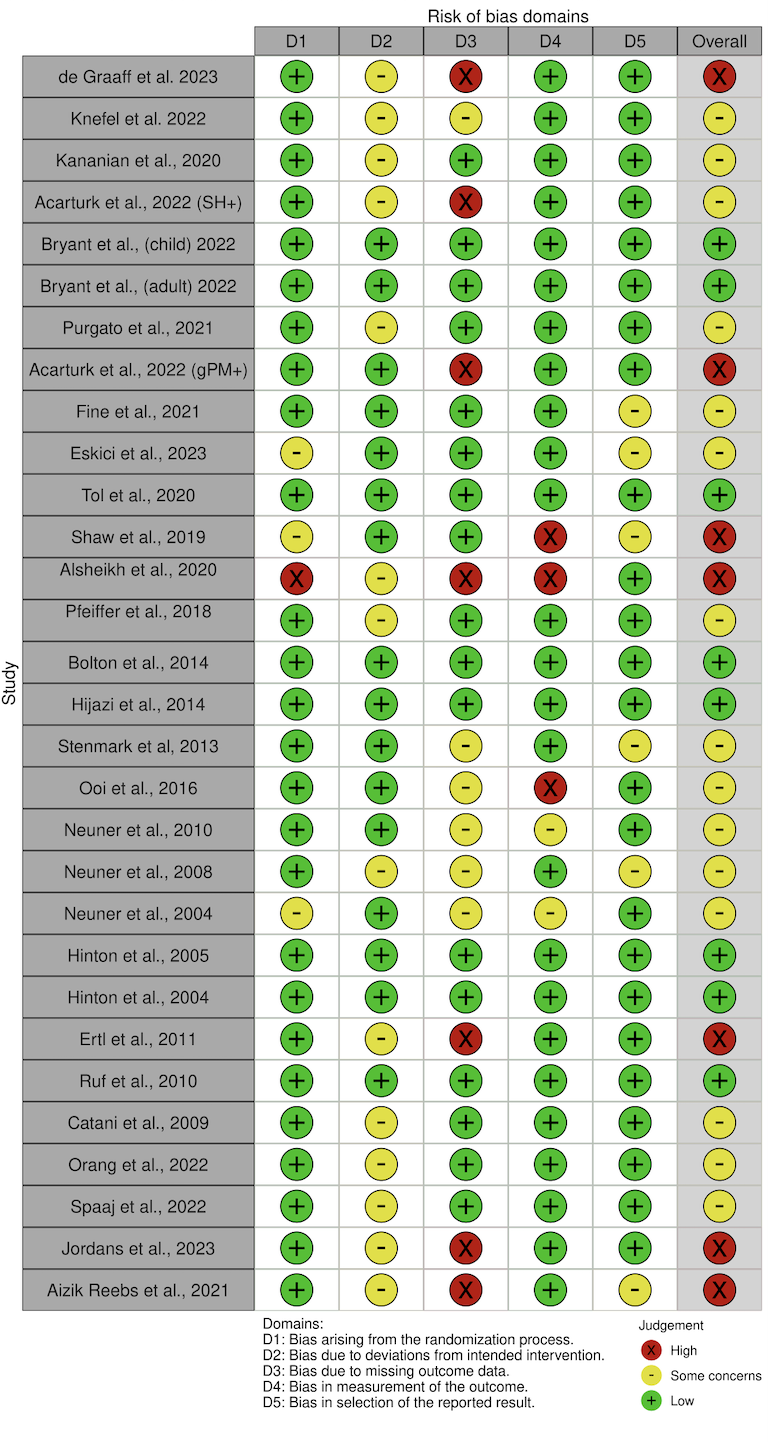

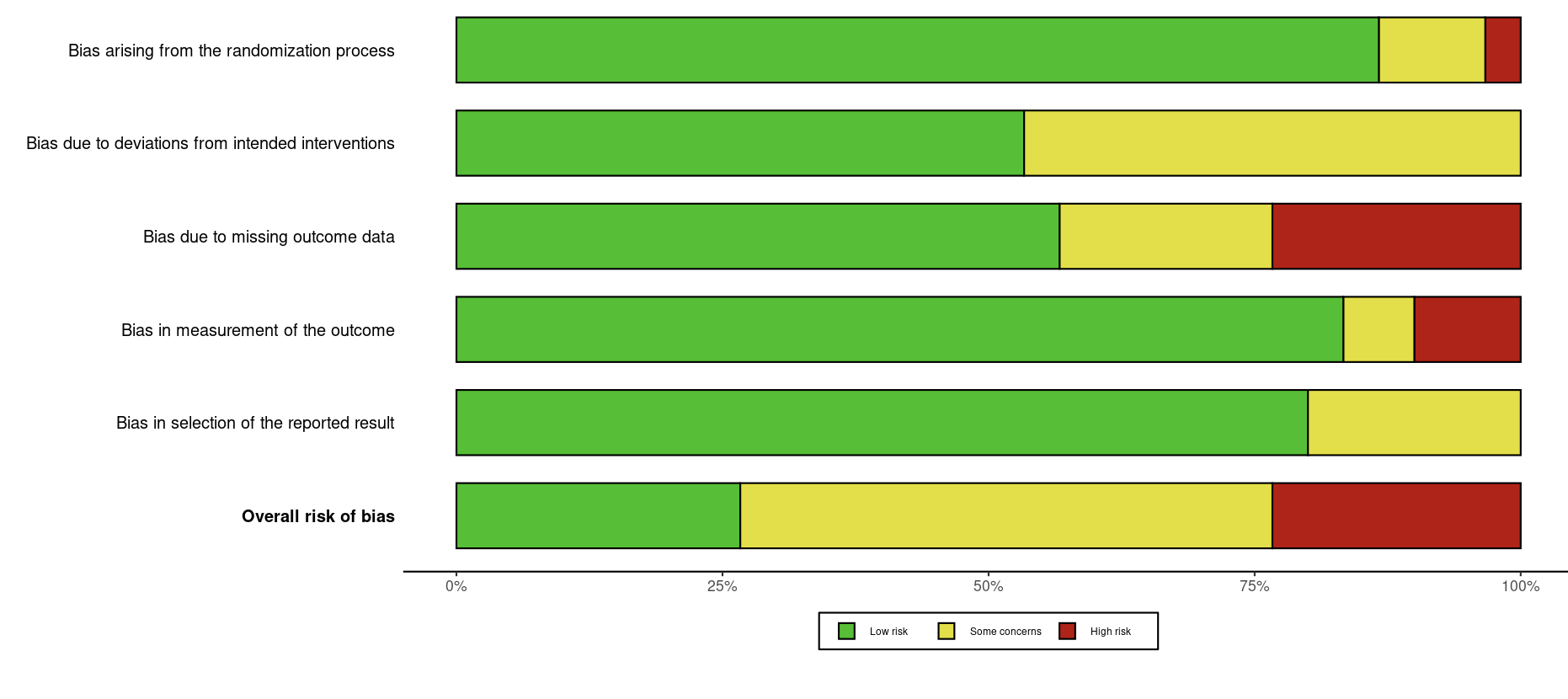

**3.1** Risk of bias summary of randomised trials: authors’ judgements about each risk of bias domain for included studies.

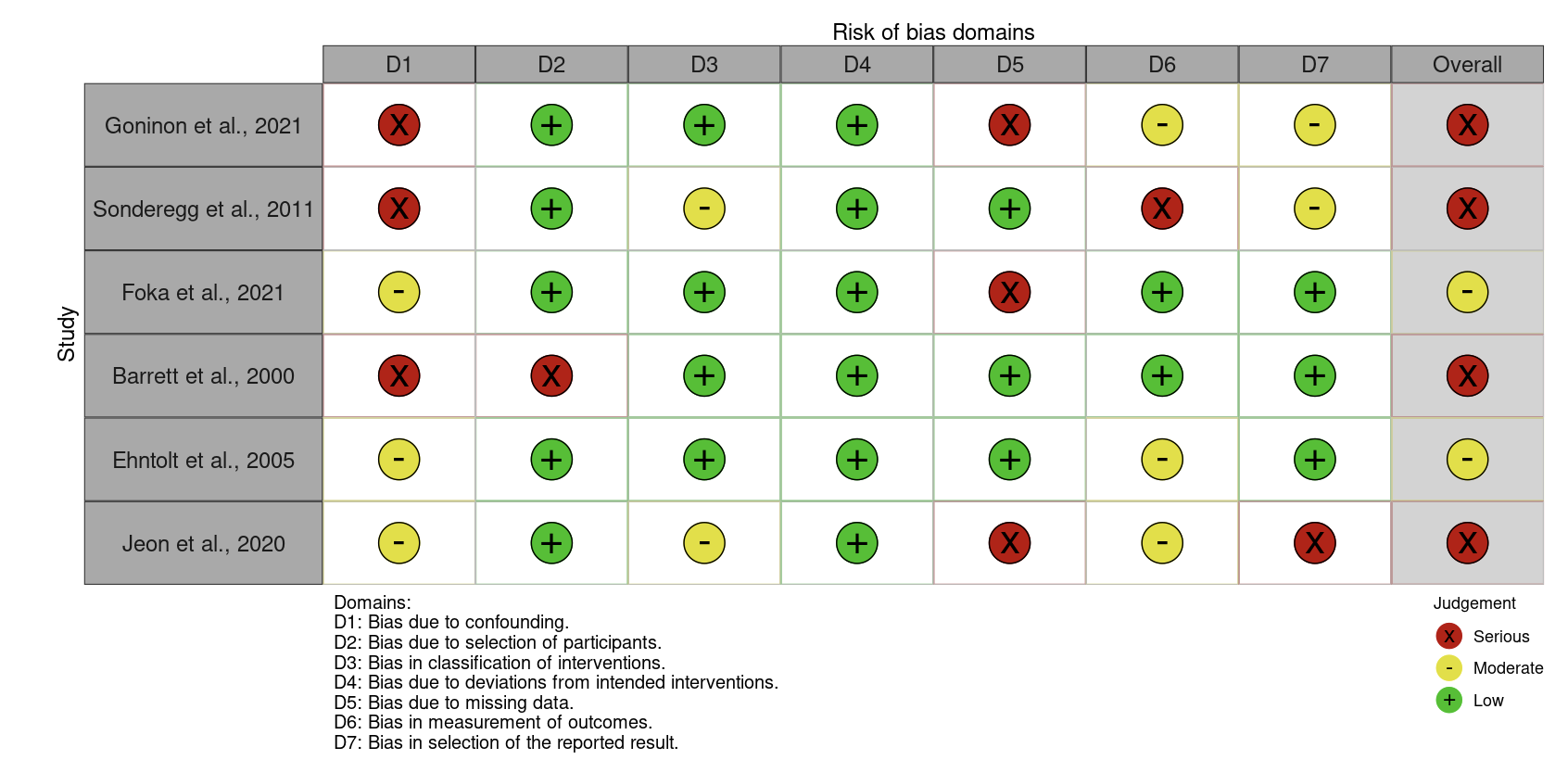

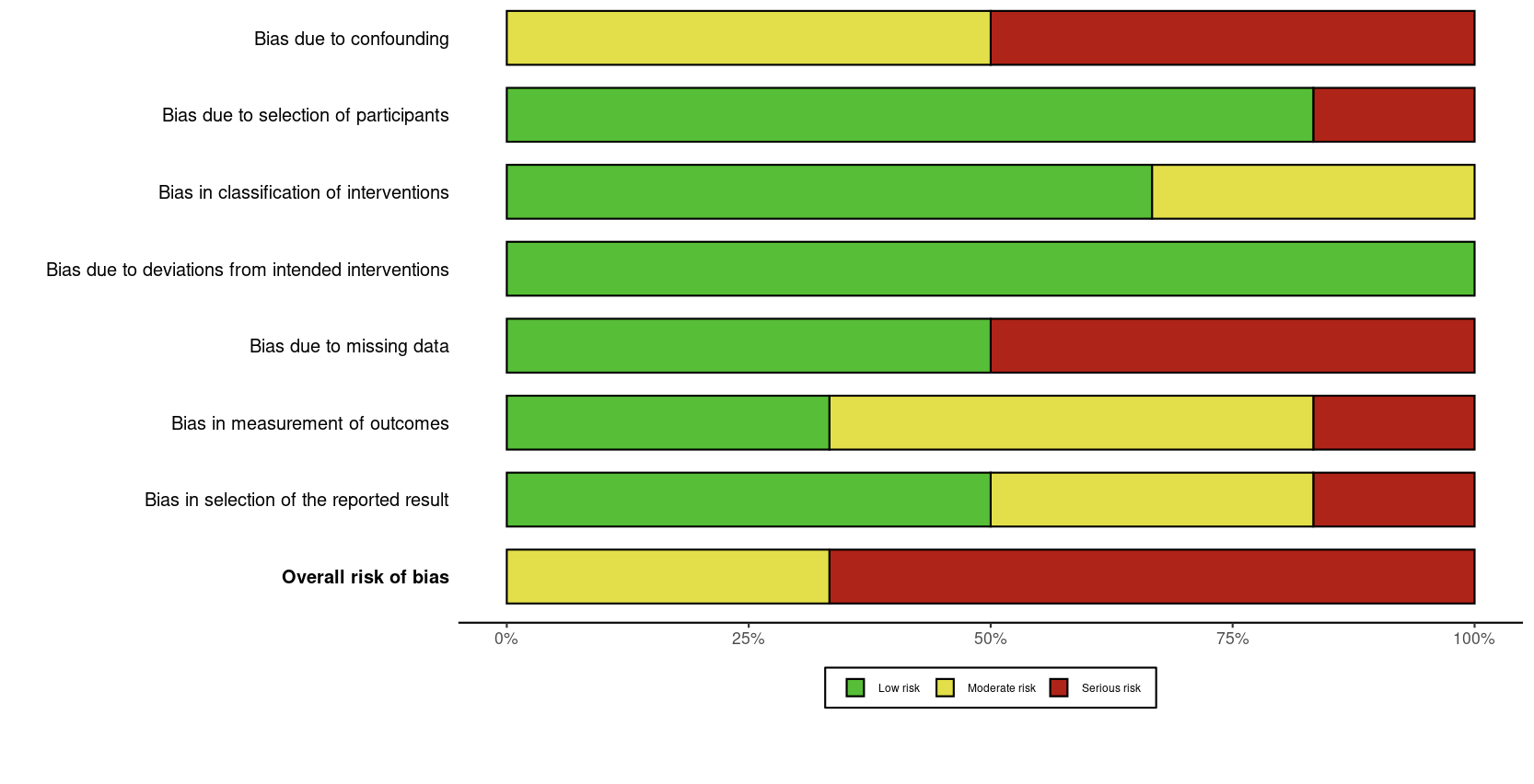

**3.2** Risk of bias summary of non-randomised trials: authors’ judgements about each risk of bias domain for included studies.

**SM4. Funnel plots**

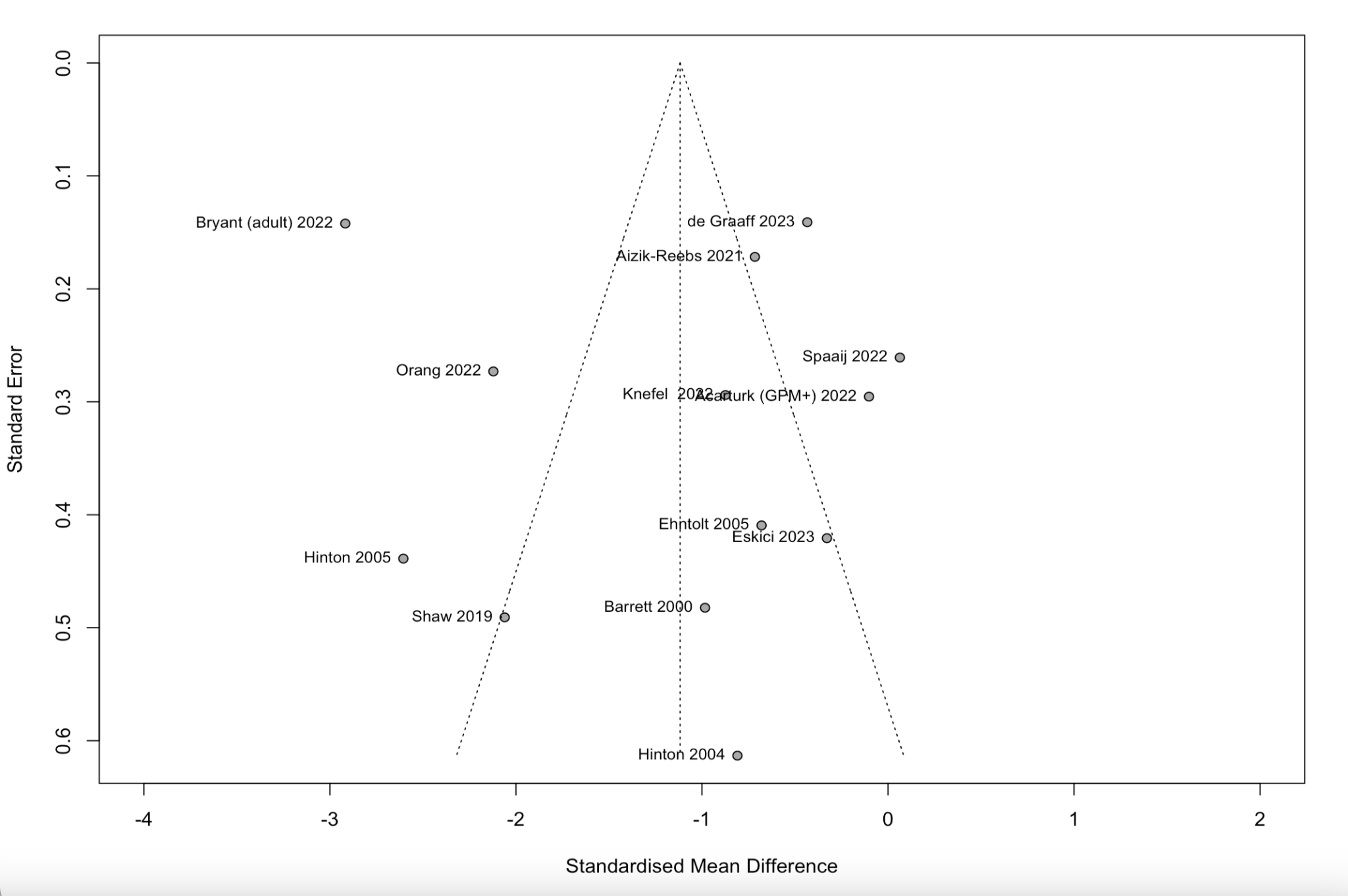

**4.1** Funnel plot of anxiety outcomes

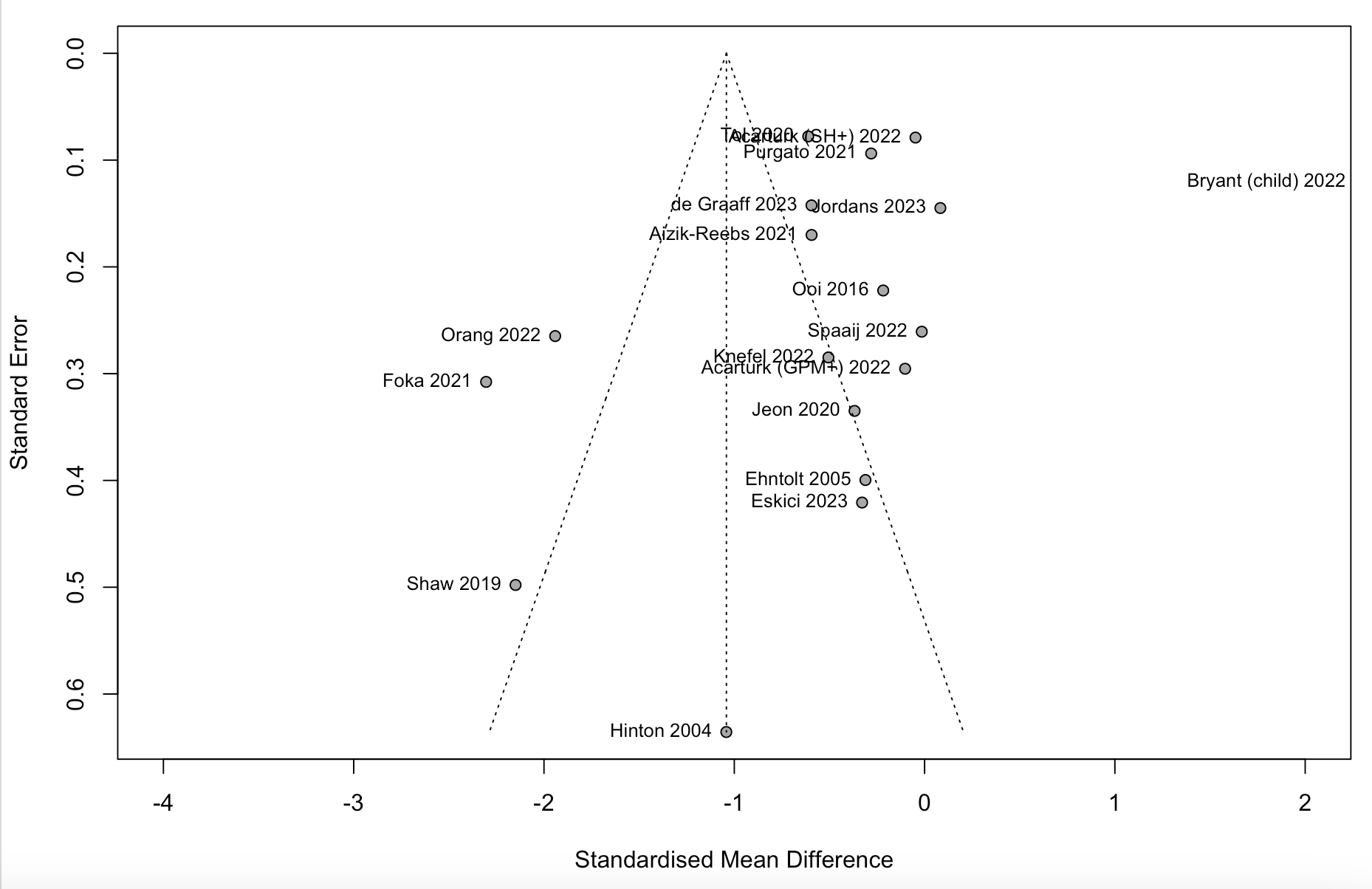

**4.2** Funnel plot of depression outcomes

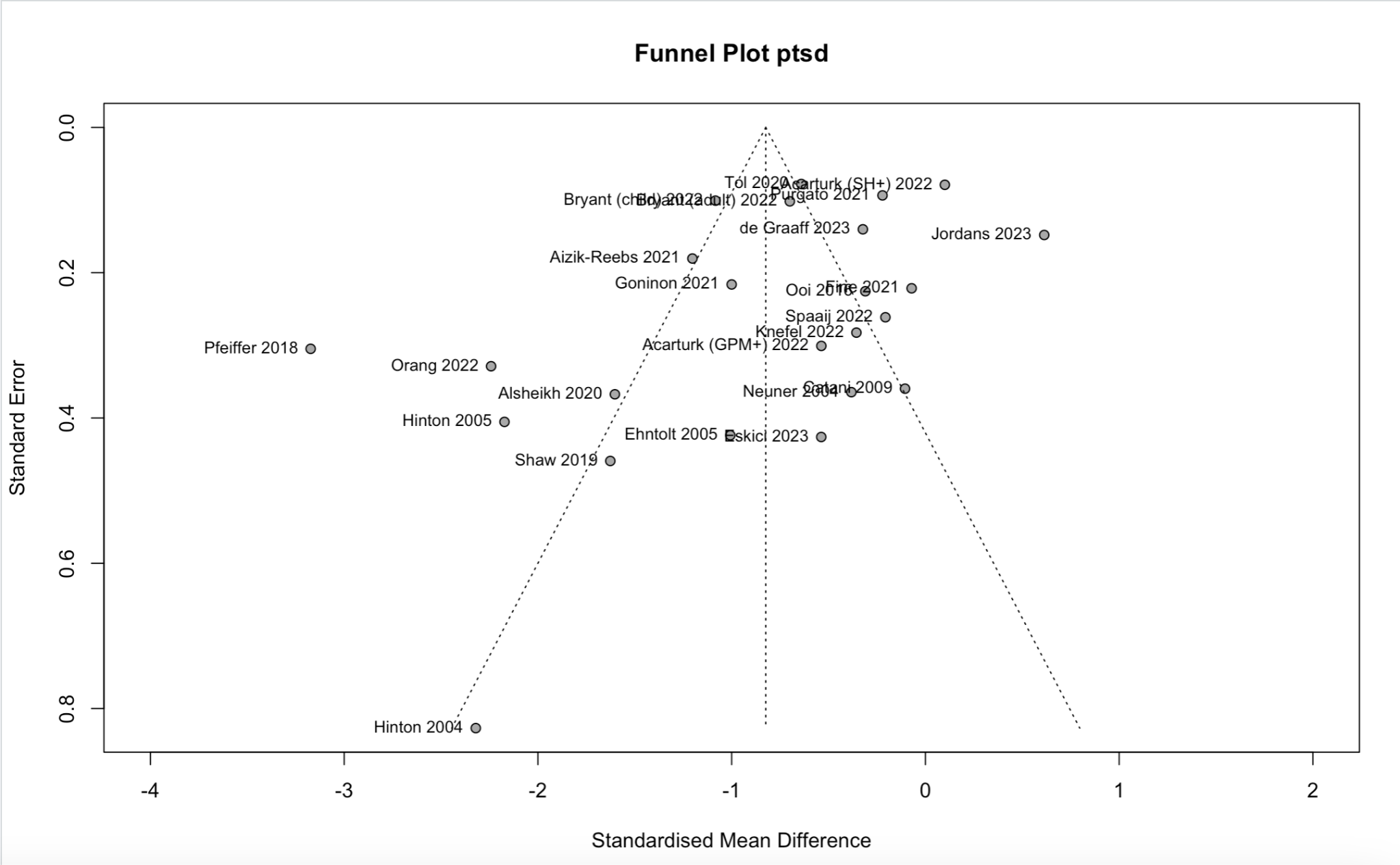

**4.3** Funnel plot of PTSD outcomes

**SM5. Subgroup analyses - LX**

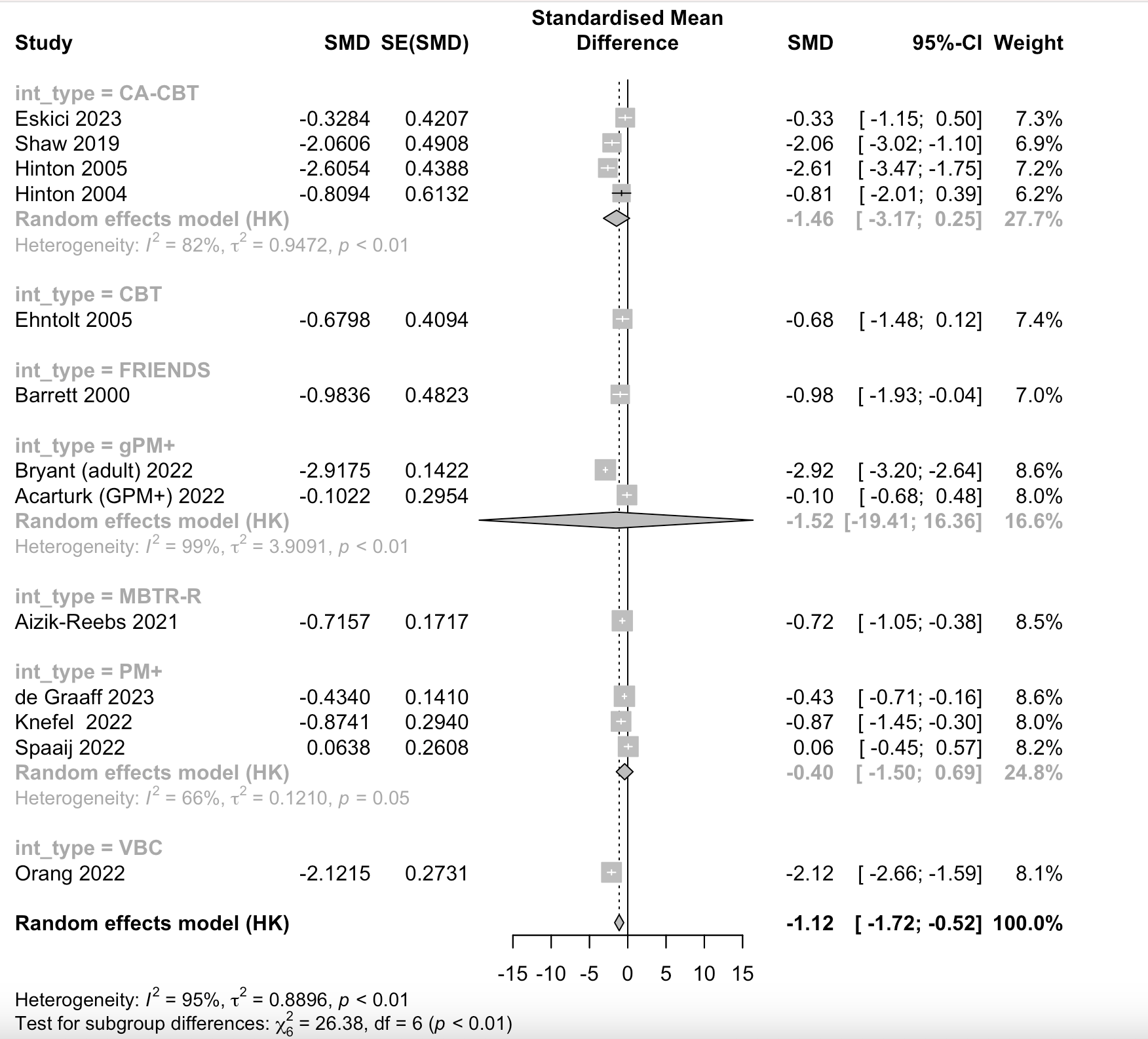

**5.1.1** Effectiveness of interventions by type of intervention method on anxiety outcomes.

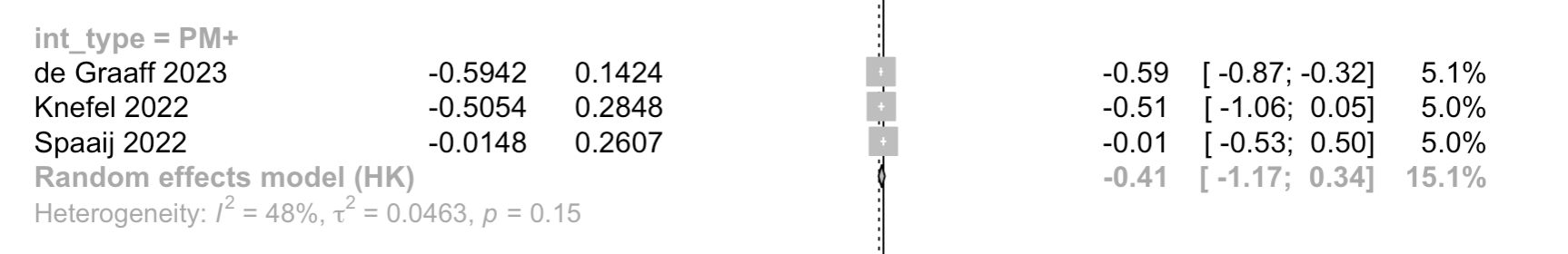

**5.1.2** Effectiveness of interventions by type of intervention method on depression outcomes.

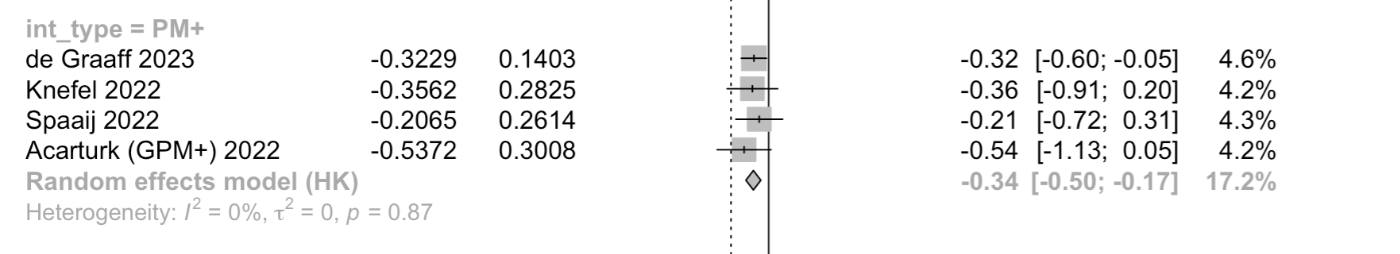

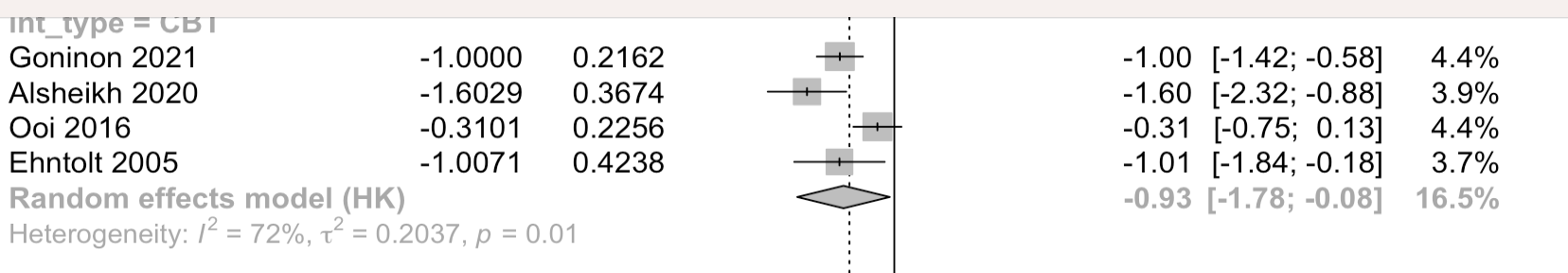

**5.1.3** Effectiveness of interventions by type of intervention method on PTSD outcomes.

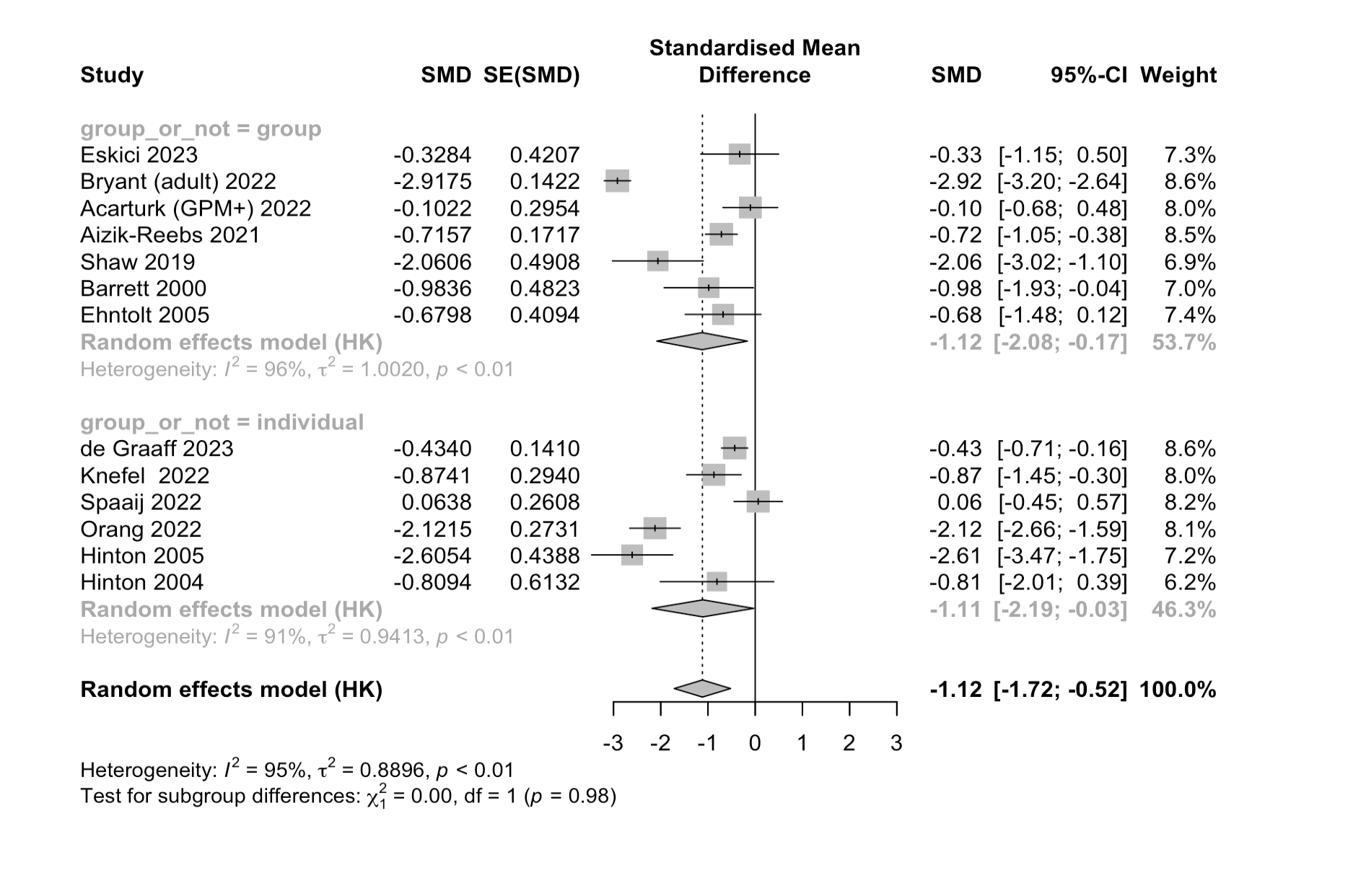

**5.2.1** Effectiveness of interventions carried out in a group setting versus individually on anxiety outcomes.

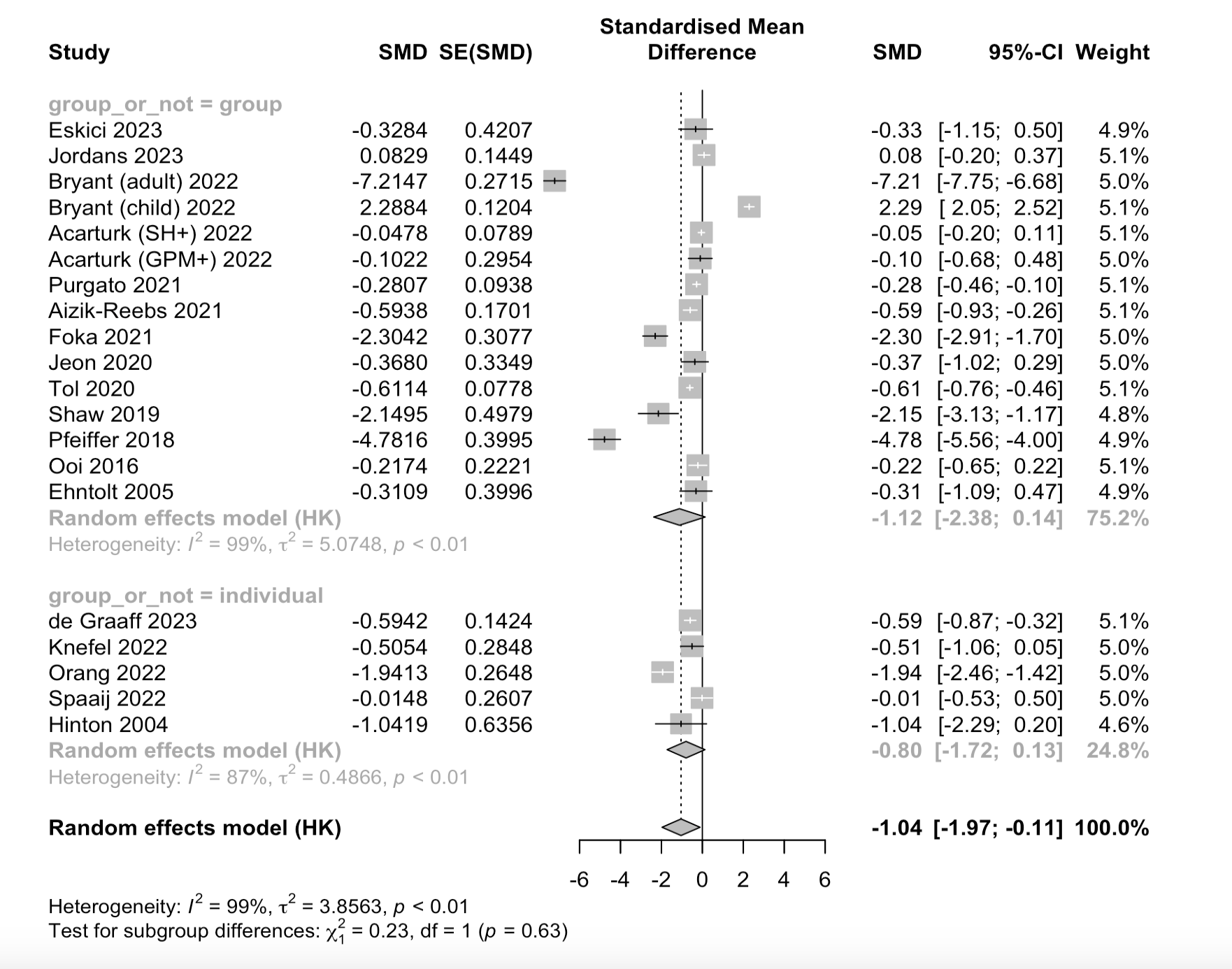

**5.2.2** Effectiveness of interventions carried out in a group setting versus individually on anxiety outcomes.

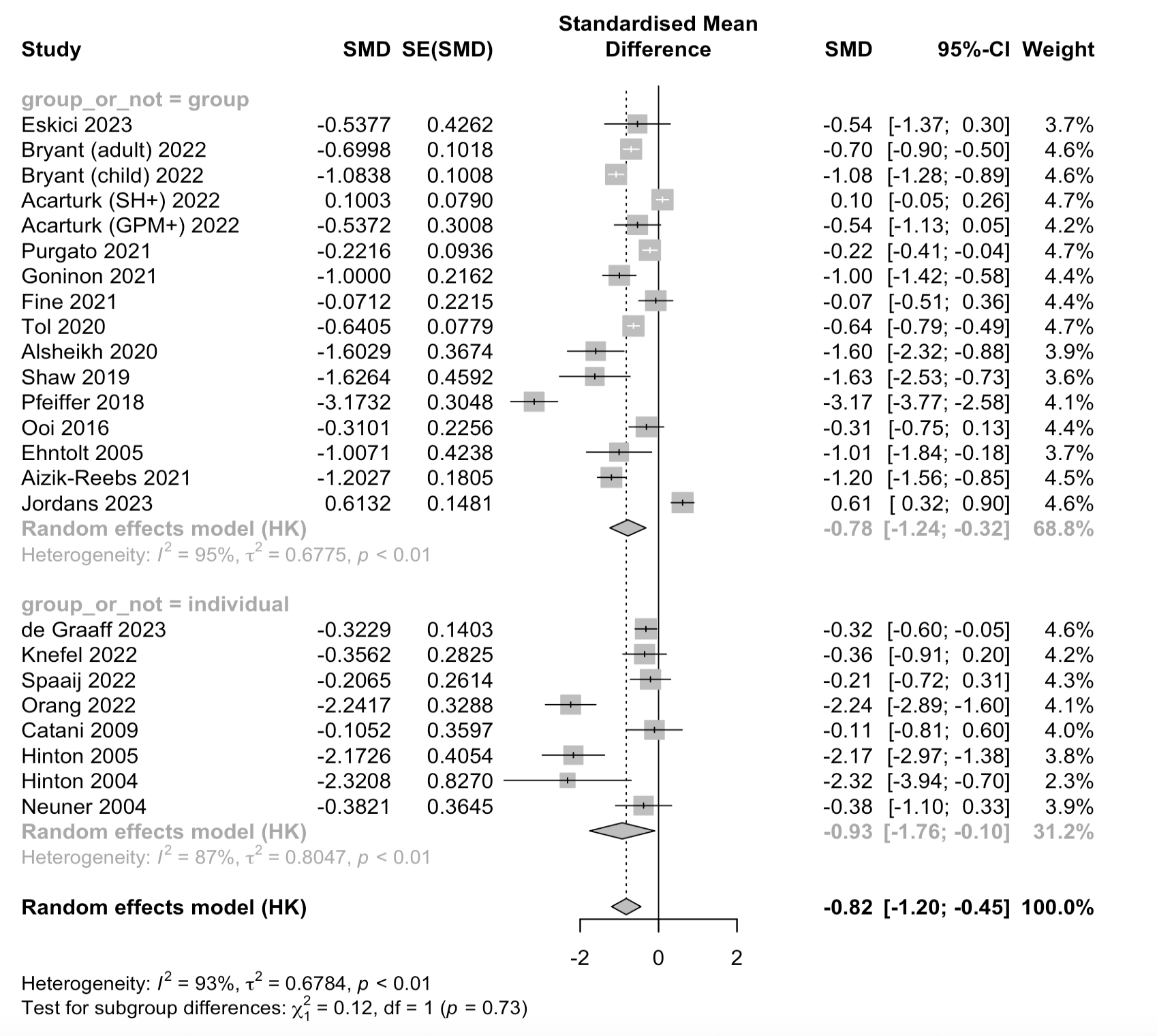

**5.2.3** Effectiveness of interventions carried out in a group setting versus individually on PTSD outcomes.

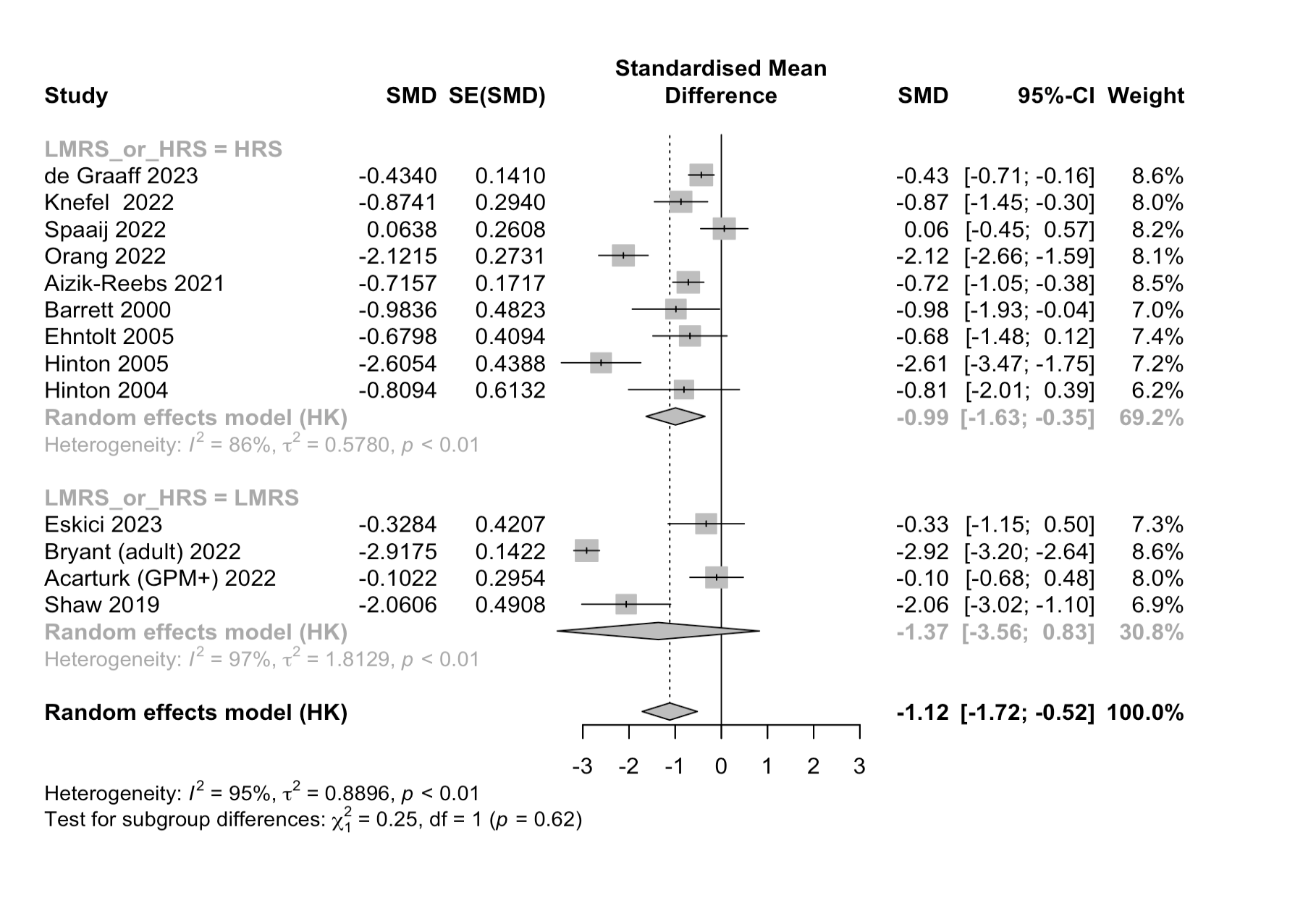

**5.3.1.** Effectiveness of interventions carried out in low-to-medium resource settings versus high-resource settings on anxiety outcomes

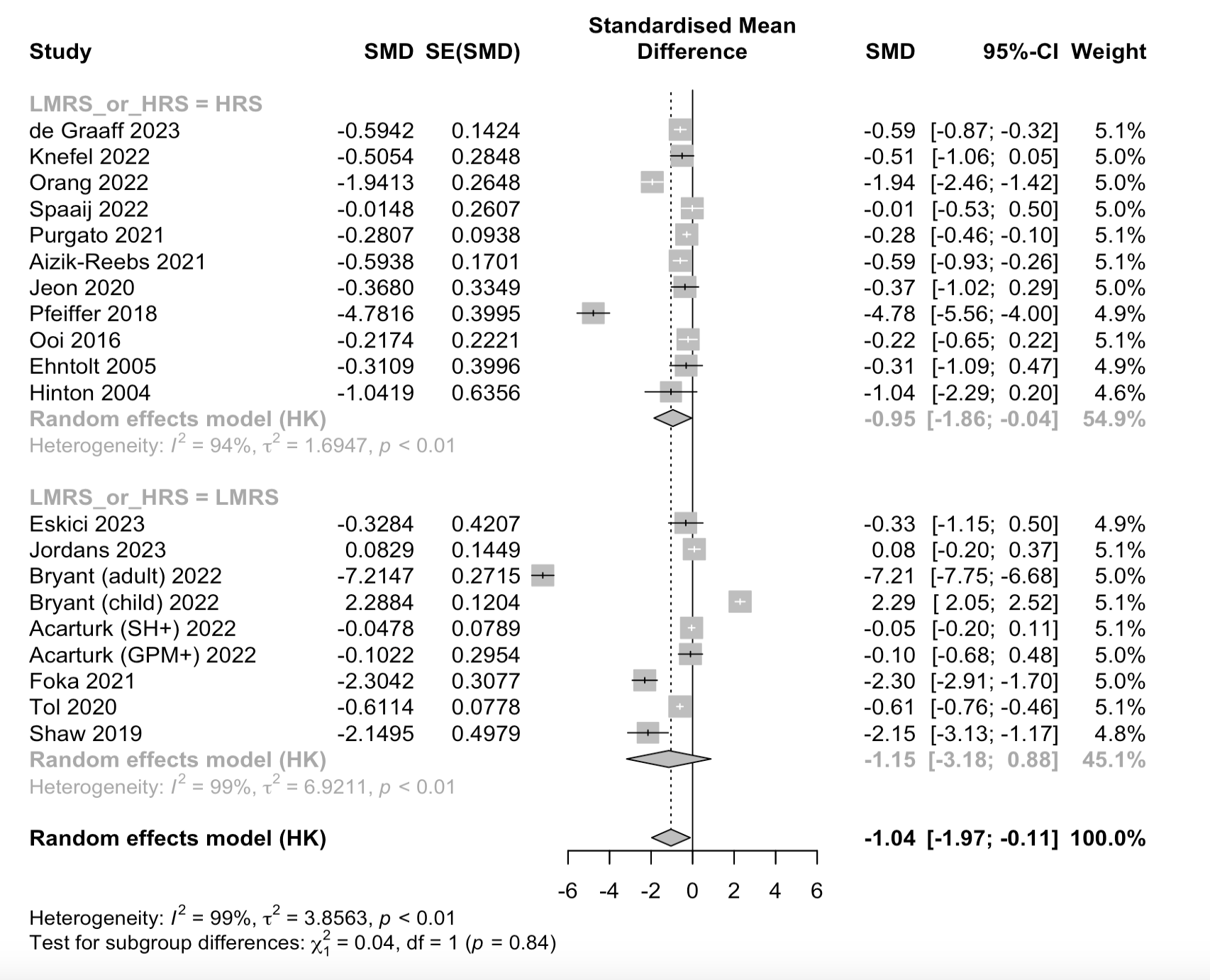

**5.3.3.** Effectiveness of interventions carried out in low-to-medium resource settings versus high-resource settings on depression outcomes.

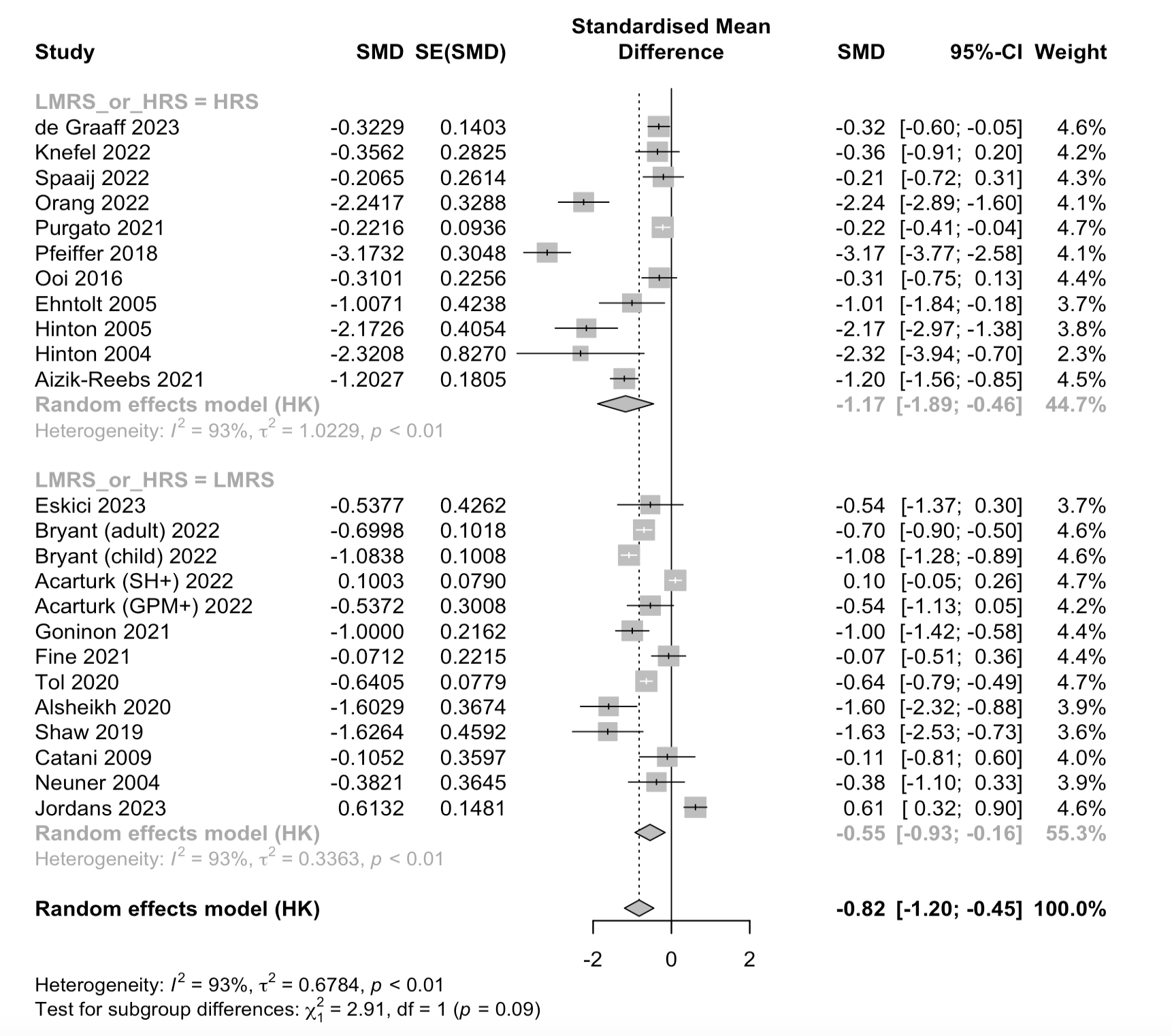

**5.3.3.** Effectiveness of interventions carried out in low-to-medium resource settings versus high-resource settings on PTSD outcomes.

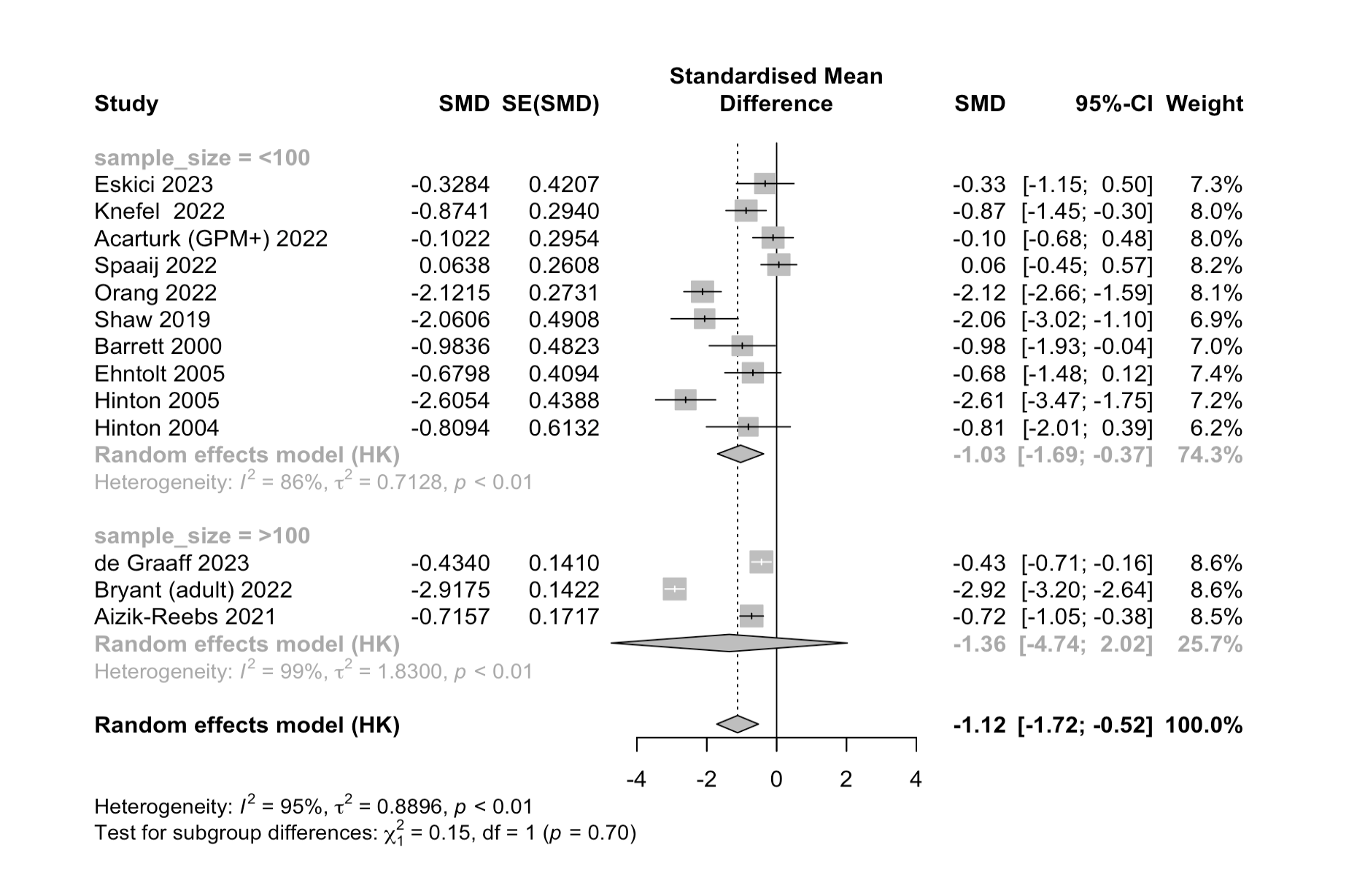

**5.4.1.** Effectiveness of interventions carried out in studies with small (*n* < 100) versus

large (*n* > 100) sample sizes on anxiety outcomes.

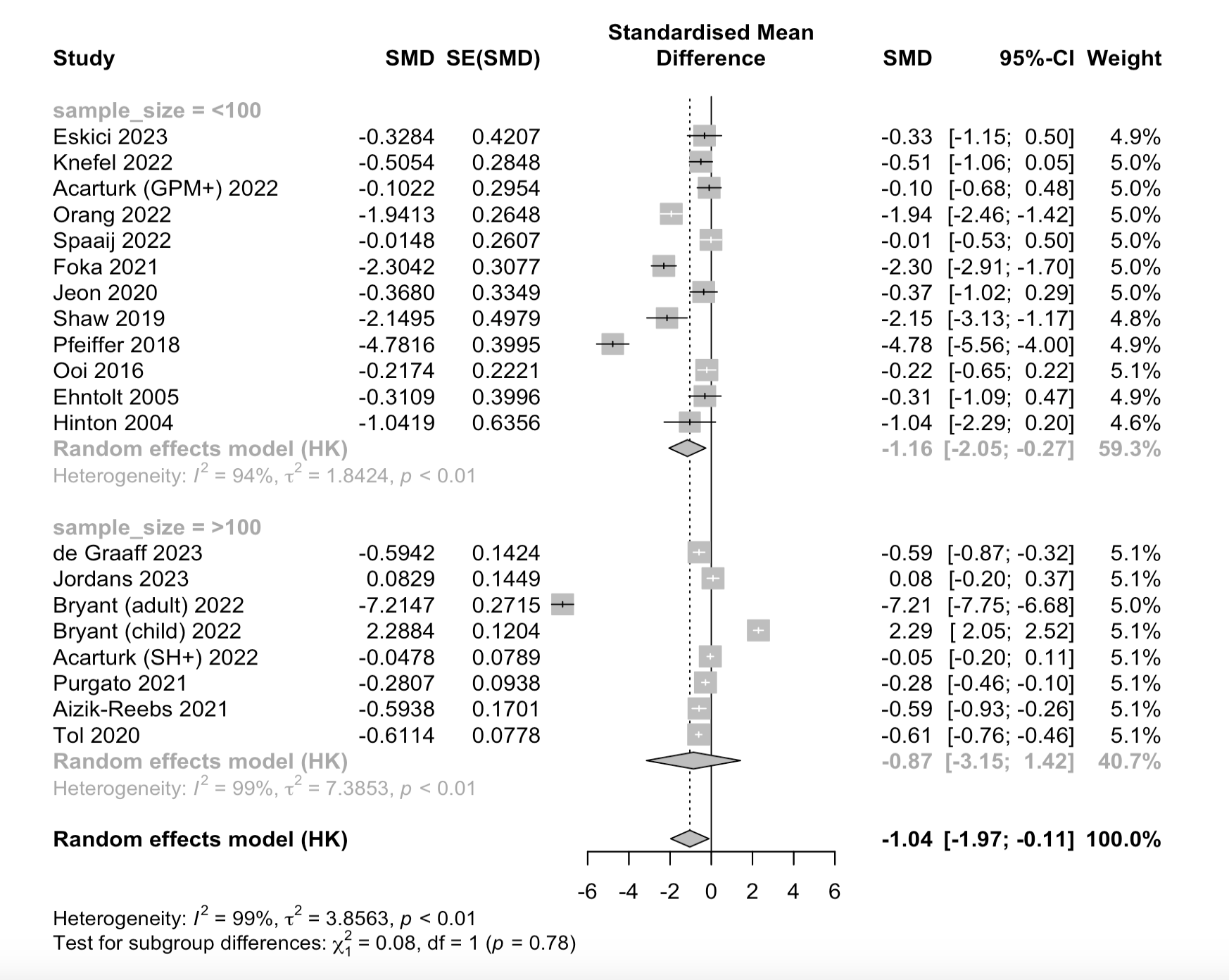

**5.4.2.** Effectiveness of interventions carried out in studies with small (*n* < 100) versus

large (*n* > 100) sample sizes on depression outcomes.

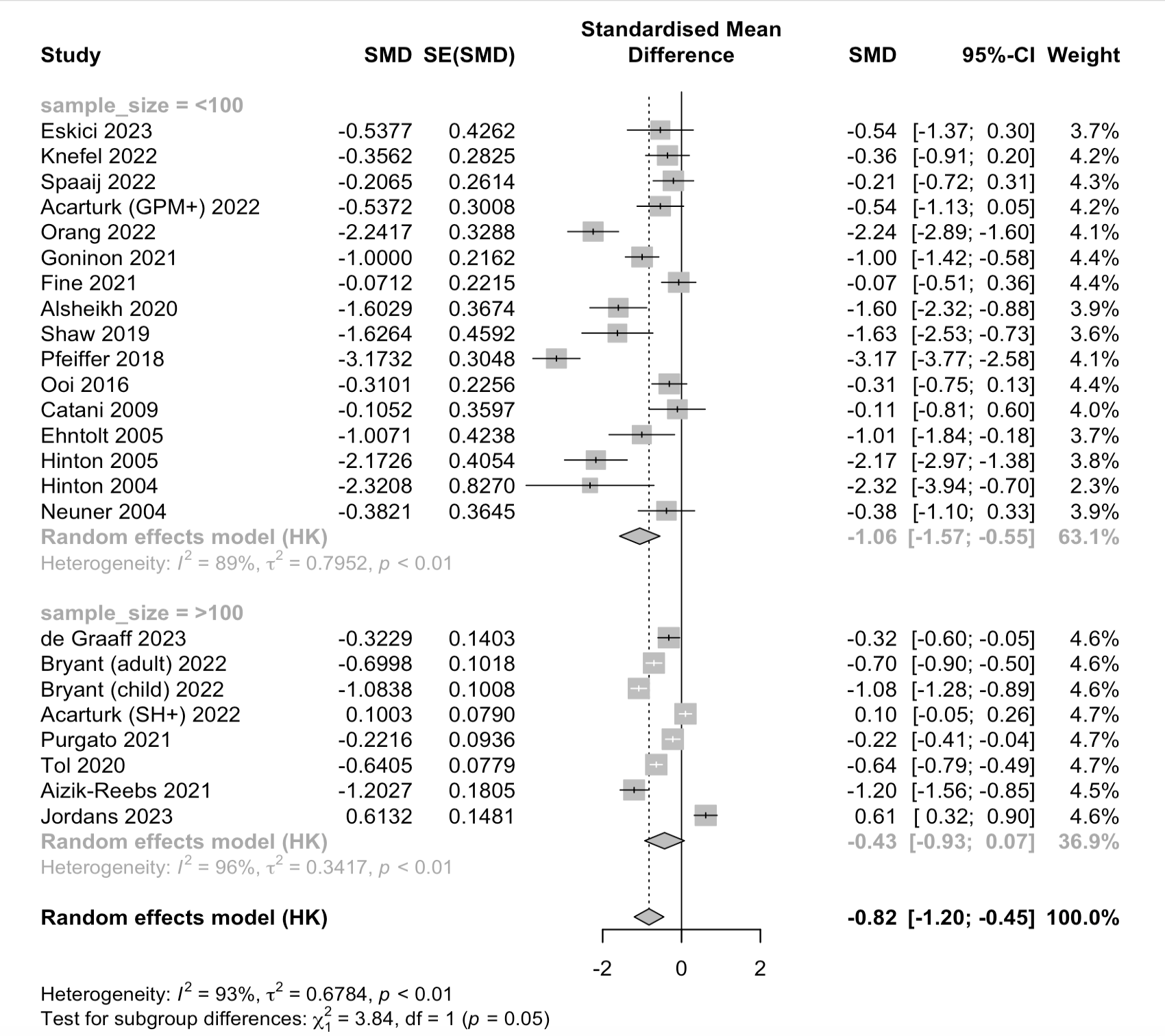

**5.4.3.** Effectiveness of interventions carried out in studies with small (*n* < 100) versus

large (*n* > 100) sample sizes on PTSD outcomes.

**SM6. Short-term follow up - ND**

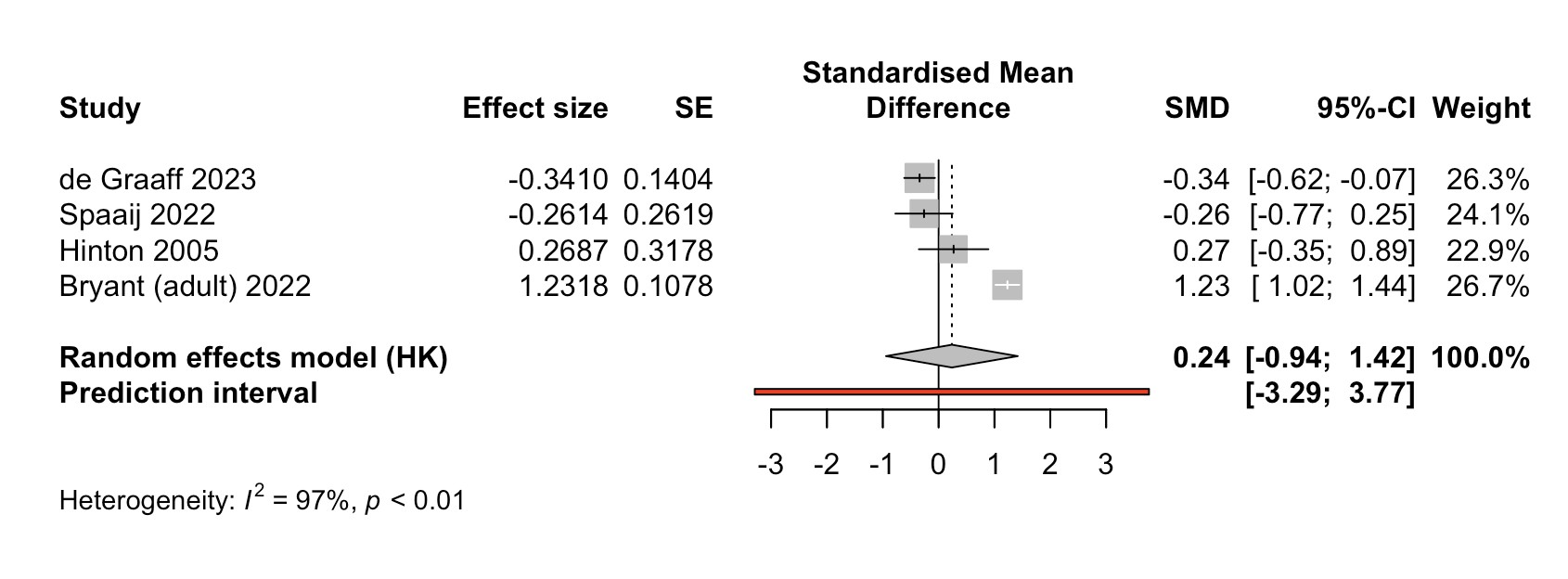

**6.1.** Effectiveness of brief CBT-based psychological interventions in treating anxiety in

refugees and asylum seekers at 3-6 month follow-up

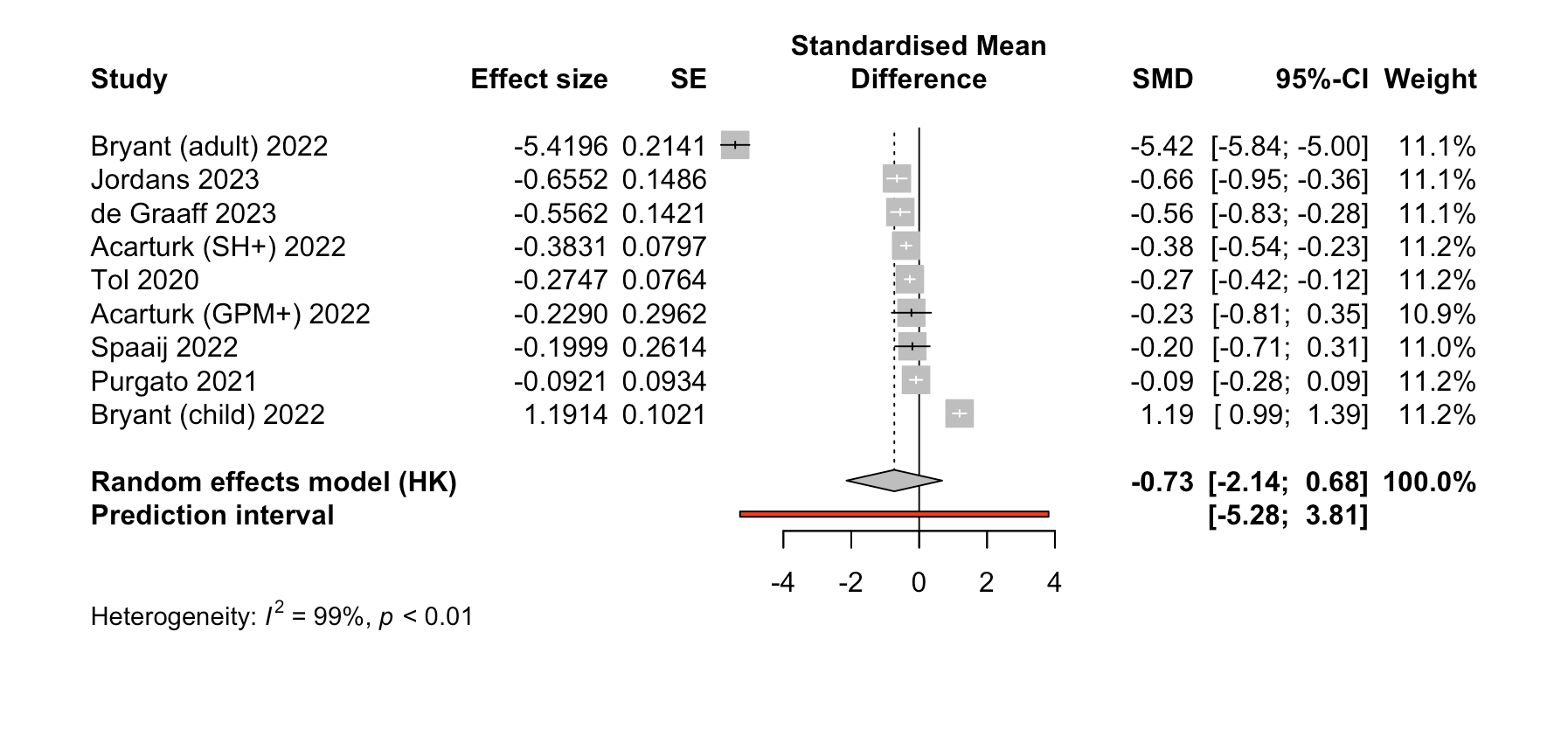

**6.2.** Effectiveness of brief CBT-based psychological interventions in treating depression in

refugees and asylum seekers at 3-6 month follow-up

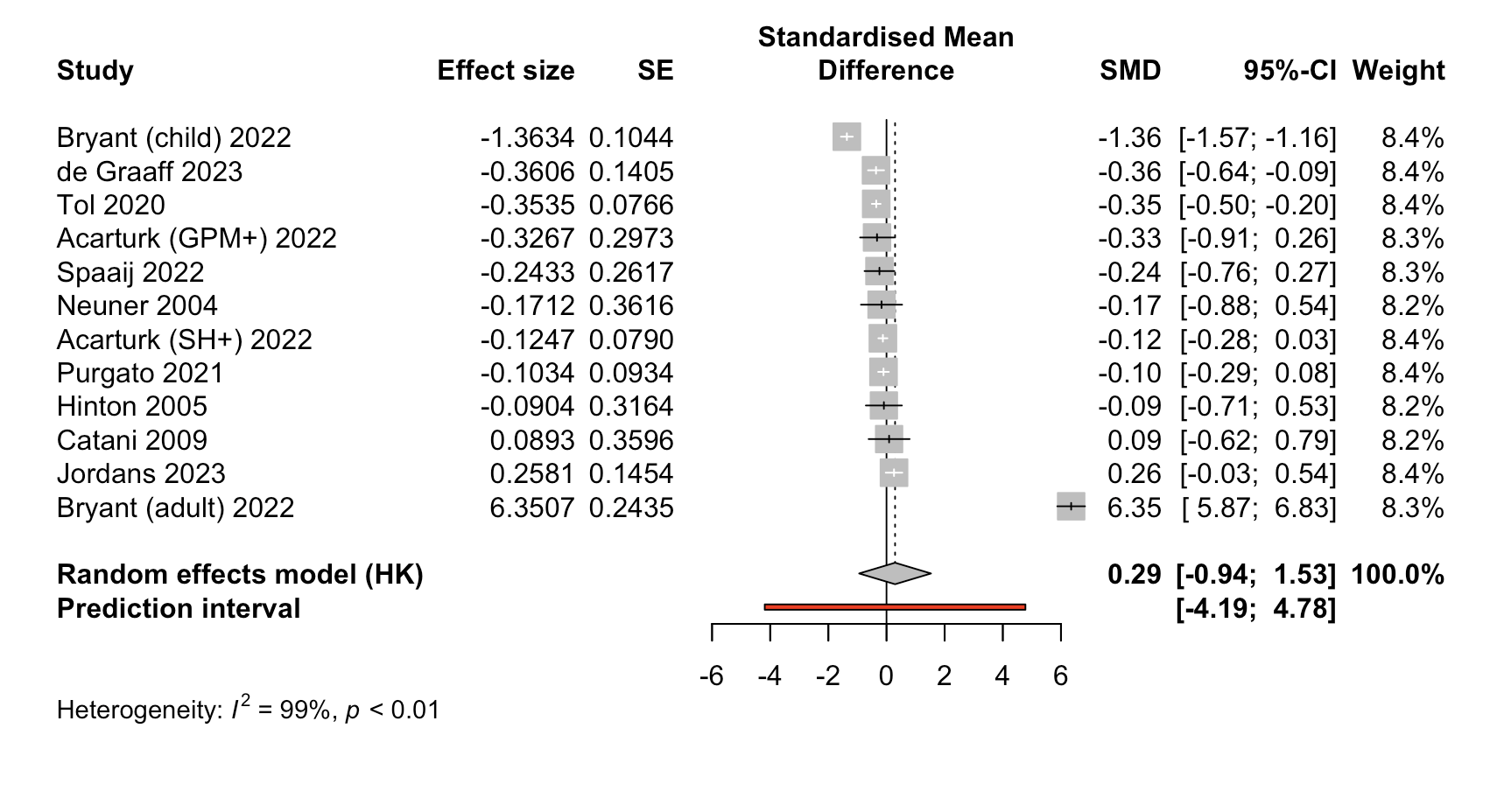

**6.3.** Effectiveness of brief CBT-based psychological interventions in treating PTSD in

refugees and asylum seekers at 3-6 month follow-up
